# Rapid health evaluation in migrant peoples in transit through Darien, Panama: Protocol for a multi-method qualitative and quantitative study

**DOI:** 10.1101/2021.10.28.21265606

**Authors:** Amanda Gabster, Monica Jhangimal, Jennifer Toller Erausquin, José Antonio Suárez, Justo Pinzón-Espinosa, Madeline Baird, Jennifer Katz, Davis Beltran-Henríquez, Gonzalo Cabezas-Talavero, Andrés F. Henao-Martínez, Carlos Franco-Paredes, Nelson I. Agudelo-Higuita., Mónica Pachar, José Anel González, Fátima Rodriguez, Juan Miguel Pascale, Migrant Peoples in Transit Study Group

## Abstract

**Background:** The world is currently unprepared to deal with a the drastic increase in global migration. There is an urgent need to develop programs to protect the well being and health of migrant peoples. Increased population movement is already evident throughout the Americas as exemplified by the rising number of migrant peoples that pass through the Darien neotropical moist broadleaf forest along the border region between Panama and Colombia. The transit of migrant peoples through this area has an increase in the last years. In 2021 an average of 9,400 people entered the region per month compared to 2,000-3,500 people monthly in 2019. Along this trail, there is no access to healthcare, food provision, potable water, or housing. To date, much of what is known about health needs and barriers to healthcare within this population is based on journalistic reports and anecdotes. There is a need for a comprehensive approach to assess the healthcare needs migrant peoples in transit. This study aims to describe demographic characteristics, mental and physical health status and needs, and experiences of host communities, and to identify opportunities to improve healthcare provision to migrant peoples in transit in Panama.

**Study design and methods:** This multi-method study will include qualitative (n=70) and quantitative (n=520) components. The qualitative component includes interviews with migrant peoples in transit, national and international non-governmental organizations and agencies based in Panama. The quantitative component is a rapid epidemiological study which includes a questionnaire and four clinical screenings: mental health, sexual and reproductive health, general and tropical medicine, and nutrition.

**Conclusion:** This study will contribute to a better understanding of the health status and needs of migrant peoples in transit through the region. Findings will be used to allocate resources and provide targeted healthcare interventions for migrant peoples in transit through Darien, Panama.

## Introduction

Currently, there are 281 million international migrant peoples worldwide, which is three times greater than the number of migrant peoples estimated in 1970 (1). Additionally, the number of forcibly displaced people worldwide has risen dramatically in the past decade (2). The world is currently unprepared to deal with population movement of this scale, and there is an urgent need to develop programs to protect the well being and health of migrant peoples. A dramatic increase in population movement is evident through the Americas, where migrant peoples from Africa, Asia, South America, and the Caribbean travel through South and Central America en route to North America. Although other portions of the route may involve travel by air, train, or bus, migrants who arrive in South America headed north must travel on foot through the Darien National Forest along the border region between Panama and Colombia.

The Darien Forest (DF), commonly called the ’Darien Gap,’ is a roadless, dense, 106 km long neotropical moist broadleaf forest that interrupts the Panamerican Highway and serves as a natural barrier between Colombia and Panama. The crossing — through what is known to be “the most dangerous and inhospitable” jungle in the world— is a treacherous journey, often taking between seven to fourteen days depending on the route taken, the amount of food carried, and travel group members’ abilities (3). Migrant peoples who travel this route have no access to potable water or food and must face deadly river crossings, scorching high temperatures, and poisonous animals. Some indigenous communities situated along the dense forest’s outskirts provide a temporary resting place for migrant peoples in transit. Drug traffickers also share the trails, and people who have made the journey have commonly reported human trafficking, smuggling and violence. Moreover, from May to September 2021, 180 cases of rape were reported to Doctors Without Borders (4).

The National Border Police (SENAFRONT) guides migrant peoples who emerge from the Gap to ’Migrant Reception Stations’ (MRS), organized by the National Migrant Services. All individuals who enter by foot into Panama, with very few exceptions, pass through these MRS. These posts are part of the international ’controlled flow’ of migrant peoples in a coordinated effort between the United States of America (USA) and Panama, where biometric measurements are taken of all individuals entering the country to screen for terrorism and keep a document trail of individuals (5, 6). Time spent at the MRS depends on several factors including the individual’s nationality, health needs and demographics. Citizens of some countries are allowed to pass quickly, while others may need to wait months for their migration process in Panama. Individuals from South America and the Caribbean (except Haiti and Cuba) are among those who have non-expedited processes and these nationals who arrive in Darien must go through lengthy months-long visa processes before continuing. Citizens from other countries often stay in the Darien MRSs for approximately 3-14 days before being transported to another MRS near the Panama-Costa Rica border, where they will remain until initiating their travels north.

From 2015 to 2021, there has been a steady increase in the number of migrant peoples in transit that pass through the DF at the Colombia-Panama border en route to North America (7–9). Prior to 2020, there were between 2,000-3,000 individuals who crossed into Panama monthly through the DF. These trails have been used for centuries to cross between the two countries, however, from 2016 onwards the number of people using this route has increased annually. From January to September 2021, more than 91,000 migrant peoples entered Panama through the DF (10). This recent increase follows a period early in the COVID-19 pandemic where a large number of people were unable to cross international borders (8, 11). Additionally, perceptions of the United States’ changes in immigration policies with the Biden administration has been identified as impacting the number of migrant peoples through these routes (12). Individuals who make this journey come from different countries (Map 1). According to official records from Panama’s National Migration Service, over 70% are Haitian citizens, 20% Cuban citizens, and the remaining 10% originate from Sub-Saharan Africa, Asia, and South America (8). Furthermore, from 2018, there has been an increase in children, families, and pregnant women entering Panama on foot through the DF (8, 13). Individuals from outside the continent arrive in South America by land or sea, often arriving in Ecuador or Brazil, beginning their journey by land up to the Panama-Colombia border (7–9).

There is no access to healthcare along the route through the DF and very limited access to needed health services while in the MRSs. During the DF crossing, migrant peoples are exposed to dangerous elements from the days spent in the tropical forest and people encountered during their route. These elements are coupled with increased prevalence of diseases and health conditions common in their countries of origin prior to beginning their route through Darien. In recent years, Panama’s Ministry of Health (MINSA) and some international organizations have provided resources to begin to address the health needs and offer healthcare to migrant peoples in transit at the MRS. However, the COVID-19 pandemic has reoriented healthcare service provision, where healthcare is focused on SARS-CoV-2 diagnosis and quarantine; however other services such as general health, sexual health, mental health, reproductive medicine and fever management have been left aside (14, 15). PAHO has recommended in their priority actions a need to strengthen surveillance and monitoring of the prevalence of health conditions through: [1] the development of comprehensive profiles of the health status of migrants, and [2] strengthening of national and decentralized health surveillance systems, especially in the border- transit areas that can capture the health status and needs of migrants (16). In accordance with these recommendations, this study protocol is intended to be a rapid assessment of migrant peoples’ health status, needs, impact on surrounding communities, and opportunities for improved service provision to this population. Results from the study will inform future healthcare for migrants in transit in Darien and the region.

## Methods

### Study design and population

This study will use both qualitative and quantitative cross-sectional methods. We will use qualitative research methods to capture the perspectives and experiences of migrant peoples in transit and stakeholders (Table 1). Additionally, we will use qualitative research to learn from the experiences of a variety of stakeholders, including those who provide services, design and allocate resources to programming, and individuals that live in the communities where migrants pass while crossing the DF. Participants within the study’s qualitative component include government health and migration personnel, representatives from international organizations working with migrant populations, community members from surrounding host communities, and migrant peoples.

**Table 1:** Study components and activities to be undertaken

|  | Qualitative (Component A) | Quantitative Component B |  |  |  |  |
| --- | --- | --- | --- | --- | --- | --- |
|  | Qualitative data collection | Quantitative questionnaire | Mental Health | Sexual and Reproductive Health | General and Tropical Medicine | Nutrition |
| Target Population | Community officials, governmental and non-governmental personnel that work with the target population, migrant peoples in transit, people that live in the communities where migrant peoples travel through or live | Migrant peoples in transit | Migrant peoples in transit | Migrant peoples in transit | Migrant peoples in transit | Migrant peoples in transit |
| Activities | Semi-structured interviews, focus groups and participant observation | Self-administered quantitative questionnaire | Clinical evaluation and screening | Clinical evaluation and screening | Clinical evaluation and screening | Clinical evaluation and screening |

The qualitative component (Component A) will use purposive sampling and undertake semi-structured interviews, focus groups and select participant observation to assess health needs and service provision to migrant peoples (Table 2). Qualitative data collection tools (i.e., semi-structured interview guides for each stakeholder group) will be designed to capture data to describe divergent health priorities and available health services, evaluate patient barriers for accessing health services, and document needed resources for improving healthcare provision according to stakeholder perspectives. Through coding and thematic analysis of interviews, results of the qualitative component will allow our research team to contextualize prevalence data from the quantitative study component. The qualitative component will provide insights from key stakeholder groups, in their own words. Interviews will provide rich descriptions of the migrant experience and health needs of migrants in transit through Darien, as well as challenges to health services provision. We will use this information to prioritize needs and identify targets and strategies for future intervention.

**Table 2:** Inclusion and exclusion criteria for the study

|  | Inclusion criteria | Exclusion criterion |
| --- | --- | --- |
| **Qualitative Study** | 1. Aged >17 years<br>2. Speak and write at least basic level one of these languages: Spanish, English, Haitian Creole, French or Portuguese | Have a planned exit from an MRS the same day the study team has reached the person |
| **Quantitative Questionnaire** | 1. Aged >17 years<br>2. Speak and write at least basic level one of these languages: Spanish, English, Haitian Creole, French or Portuguese |  |
| **Mental health** | 1. Aged >12 years<br>2. Speak and write at least basic level one of these languages: Spanish, English, Haitian Creole, French or Portuguese |  |
| **Sexual and Reproductive Health** | 1. Aged >12 years<br>2. Speak and write at least basic level one of these languages: Spanish, English, Haitian Creole, French or Portuguese |  |
| **General and Tropical Medicine** | 1. Speak and write at least basic level one of these languages: Spanish, English, Haitian Creole, French or Portuguese |  |
| **Nutrition** | 1. Speak and write, or have a guardian if under 12 that speaks and writes at least basic level one of these languages: Spanish, English, Haitian Creole, French or Portuguese |  |

The quantitative component (Component B, Table 1) includes a rapid epidemiological study among migrant peoples in transit. This component will include a self-administered questionnaire, and four clinical screenings: mental health, sexual and reproductive health, general and tropical medicine, and nutrition. Age of inclusion for each subcomponent can be found in Table 2. The self-administered questionnaire (Supplementary material A) includes demographics, migration routes, sexual and reproductive health and behaviours and self-reported health. This questionnaire was developed from existing instruments (some modified for this population) (17–26). For the clinical laboratory components, participants will be asked to provide a mid-stream urine sample and a peripheral blood sample for laboratory point-of-care screenings for the following infections/conditions: hepatitis A,B,C and E, Chagas disease, malaria, dengue, chikungunya, HIV, syphilis and pregnancy (Table 3). Further rapid testing will be applied, according to clinical presentations, as indicated in Table 4. Clinical screening will include a review of the present and past medical history, vital signs, body measurements and a physical examination. On-site treatment (Table 5) and/or referral to primary, secondary or teritiary centers will be organized (supplementary material B) according to clinical and laboratory findings.

**Table 3:** Study activity, target population, and primary outcomes

|  | **Activity** | **Target population** | **Site** | **Primary outcomes** |
| --- | --- | --- | --- | --- |
| **Qualitative Component** | Focus groups | Host communities | Host communities | Effects of migrant presence on communities, health priorities, and needs. |
|  | Semi-structured interviews and participant observation | Migrant peoples in transit | MRS | Migration experience, barriers to health access and existing programs, health needs |
|  | Semi-structured interviews and participant observation | Public institution and non-profit national/international organization leaders who work with migrant populations in transit | MRS, health centers, institutional and organization offices | Health services available, barriers to access, needs of institutions and organizations |
| **Quantitative Component** | Self-administered questionnaire | Migrant peoples in transit | MRS | • Sociodemographic and journey information<br>• Human smuggling and trafficking<br>• Education and employment<br>• Self-rated health status<br>• Sexual biography and sexual experience<br>• Female genital cutting (for some countries)<br>• Pregnancy and fertility<br>• Self-reported STI/HIV current and past<br>• Non-consensual sex<br>• Whereabouts of family members and communication with family<br>• Felt social support |
|  | Mental health | Migrant peoples in transit | MRS | • Use of Refugee Health Screener to do an initial screen of PTSD, depression, anxiety, and somatization.<br>• MINI Neuropsychiatric International Interview: Diagnosis by DSM-5 and ICD-10 criteria<br>• EUROHIS Quality of life interview: subjective quality of life |
|  | Sexual and reproductive health | Migrant peoples in transit | MRS | • Screening for vaginal and urethral discharge disease<br>• Screening for genital ulcer disease<br>• Urine dipstick test (leukocyte esterase and nitrite) |
|  |  |  |  | • HIV rapid test (4<sup>th</sup> generation)<br>• Syphilis rapid test, RPR titer confirmatory |
|  | General and tropical medicine | Migrant peoples in transit | MRS | • Rapid point of care tests for:<br>○ HAV<br>○ HBV<br>○ HCV<br>○ HEV<br>○ Chagas<br>○ Malaria<br>○ Dengue<br>○ Chikungunya<br>○ Pregnancy<br>○ Further screening based on clinical presentation (Table 4)<br>• Clinical screening:<br>○ Review of present and past medical history<br>○ Vital signs: Blood pressure, heart rate, respiratory frequency, temperature, blood oxygen saturation<br>○ Physical examination based on systems: neurological, cardiovascular, respiratory, gastrointestinal, hepato-splenic, skin, and genitourinary |
|  | Nutrition | Migrant peoples in transit | MRS | • Screening for malnutrition and undernourishment<br>○ Height-weight<br>○ Body Mass Index (BMI)<br>○ MUAC<br>○ Marasmus and Kwashiorkor screening<br>○ MUST<br>○ Skinfold test<br>○ OMS growth charts |

**Table 4:** Further test based on clinical criteria for inclusion

| *Test* | *Clinical Criteria for inclusion* |
| --- | --- |
| COVID-19 (Antigen or PCR) | Fever or clinical suspicion |
| Crypto-Giardia-Entamoeba (antigen) | Diarrhea |
| Cytomegalovirus (antibody) | HIV positive, pregnant women with clinical suspicion |
| Yellow Fever (PCR) | Fever or clinical suspicion |
| Amsel Criteria | Vaginal secretion and suspicion of candidiasis |
| *Trypanosoma cruzi* detection (Blood smear) | Chagas positive test result, presence of acute symptoms, and suspicion based on country of origin |
| Leishmaniasis (Tissue Giemsa Stain) | Suggestive ulcer and clinical suspicion |
| *Mycobacterium leprae* (Ziehl-Nielsen Stain or Fite-Faraco Stain) | Clinical suspicion of hypochromic anesthetic lesion or nodule |
| Leptospira (serum IgGIgM antibody) | Fever or clinical suspicion |
| Malaria- (thick blood smear) | Clinical suspicion and based on country of origin (if endemic) |
| Rota/adenovirus (Stool antigen) | Pediatric age with diarrhea or gastroenteritis, in adults: fever and symptomatic respiratory |
| Measles (antibody) | Fever, malaise rash, cough, coryza, conjunctivitis, enanthem, and morbilliform exanthem |
| TORCH: Toxoplasmosis, Rubeola, CMV, HSV 1 and 2 (antibody) | Pregnant and in 1-month-old babies |
| Toxoplasmosis (antibody IgG) | HIV positive, pregnancy, and/or clinical suspicion |
| Tuberculosis (Sputum Bacilloscopic) | Fever, cough, nocturnal sweat, HIV + |
| Typhus / Para-typhus (Blood antigen) | Fever and clinical suspicion |

**Table 5:** On-site treatment by study subcomponent

| Study sub-component | Treatment Given On-Site for Diagnosed Etiology or Based on Clinical and Syndromic Suspicions |
| --- | --- |
| **Mental Health** |  |
|  | Distress, psychosis, and/or risk of self or hetero-aggressive outburst: Management with verbal containment and, if needed, pharmacologic management with 5mg of diazepam, repeated up to two times; if not, olanzapine 5mg PO or haloperidol 5mg IM. |
| **Sexual and Reproductive Health** |  |
|  | 1. Genital Herpes simplex: Acyclovir 400mg PO TID for seven days.<br>2. Recurrent genital Herpes Simplex: Acyclovir 800mg PO TID for 2 days or 400mg PO TID for 5 days.<br>3. Syphilis: Benzathine Penicillin 2.4 million UI single-dose IM , once or each consecutive week for three weeks depending on patient history/RPR<br>4. Syphilis in pregnant women: 2.4 million IU IM each week for 3 weeks<br>5. Chancroid: Azithromycin 1g PO single dose OR Ceftriaxone 250mg single dose IM OR Ciprofloxacin 500mg bid for 3 days<br>6. Chancroid in pregnant/lactating women: Erythromycin 500mg PO QID for 15 days<br>7. *Lymphogranuloma venerum*: Doxycycline 100mg PO daily for 21 days OR Erythromycin 500mg PO QID for 21 days OR Azithromycin 1g PO each week for 3 weeks<br>8. *Neisseria gonorrhoeae*: Ceftriaxone 500mg IM single dose<br>9. *Chlamydia trachomatis*: Azithromycin 1g PO single dose or Doxycycline 100mg PO bid for 7 days.<br>10. *Trichomonas vaginalis*: Metronidazole 2g PO single dose<br>11. *Candida albicans*: Fluconazole 150mg PO single dose OR Clotrimazole ovules 100mg intravaginal for 7 days<br>12. Bacterial vaginosis: Metronidazole 2g PO single dose |
| **General and Tropical Medicine** |  |
|  | 1. Cardiovascular system: Symptoms associated with myocarditis, endocarditis, pericarditis with clinical suspicion of:<br>1.1 Mild leptospirosis: Doxycycline 100mg PO bid for 7 days OR Azithromycin 500mg PO each day for 3 days<br>1.2 *Trypanosoma cruzi*: Benznidazole 5-7mg/kg/day PO divided in two doses: bid for 60 days<br>1.3 *Taenia solium*: Praziquantel 10mg/kg PO single dose<br>1.4 *Ascaris lumbricoides*: 400mg PO single dose<br>1.5 *Strongyloides stercoralis*: Ivermectin 200 µg/kg/day PO for 1- 2 days<br>1.6 *Paragonimus westermanii*: Praziquantel 25mg/kg PO tid for 2 days<br>1.7 *Mansonella perstans*: Doxycycline 200mg PO each day or 100mg PO bid for 6 weeks<br>2. Congestive Heart Failure symptoms that might be due to: |
|  | 2.1 *Schistosoma mansonii*: Praziquantel 40mg/kg PO single dose<br>3. Loeffler Syndrome that might be due to:<br>3.1 *Ascaris lumbricoides*: Albendazole 400mg PO single Dose<br>3.2 *Strongyloides stercoralis*: Ivermectin 200 µg/kg/day PO for 1- 2 days<br>3.3 *Ancyclostoma duodenale*: Albendazole 400mg PO for 1-3 days<br>3.4 *Necator americanus*: Albendazole 400mg PO for 1-3 days |
|  | **4. Dyspnea or Pneumonia suspicion due to:**<br>4.1 *Streptococcus pneumoniae*:<br>- With recent use of antibiotics and comorbidities: Amoxicillin-Clavulanate 875/125mg- 1 tablet PO bid + Clarithromycin 500mg PO bid for 7 days OR Azithromycin 500mg PO in the first dose and then 250mg PO each day for 4 days<br>- With recent use of antibiotics and no comorbidities: Amoxicillin 1g PO bid for 7 days<br>- -No recent use of antibiotics: Clarithromycin 500mg PO bid for 7 days or Azithromycin 500mg PO in the first dose and then 250mg PO each day for 4 days.<br>4.2 *Mycoplasma pneumoniae* / *Chlamydia pneumoniae* / *Chlamydia psittaci*: Doxycycline 100mg PO bid for 7 days<br>4.3 *Haemophilus influenzae*: Amoxicillin/Clavulanate 875/125mg tab one tablet PO bid for 7 days.<br>4.4 Legionellosis: Levofloxacin 750mg PO each day for 10 days<br>4.5 Coccidioidomycosis: Fluconazole 400mg PO each day for 3-12 months<br>4.6 Blastomycosis: Itraconazole 200mg PO tid for 3 days and then each day for 6-12 months<br>4.7 *Schistosoma mansonii*: Praziquantel 40mg/kg PO single dose<br>4.8 Filariasis: Doxycycline 100mg PO each day for 6-8 weeks |
|  | 5. Gastrointestinal symptoms- Hepato-splenomegaly by clinical diagnosis or suspicion of the following etiologies:<br>5.1 *Schistosoma mansonii* / *japonicum*: Praziquantel 40mg/kg-60mg/kg PO single dose (without previous treatment, dependent on country of origin) Suspiacian of acute toxemic shistosomiasis: short corse high-dose prednisone + 40mg/kg-60mg/kg Praziquantel PO in single dose, repeat Praziquantel in 4-6 weeks.<br>5.2 Toxocariosis: Albendazole 400mg PO bid for 5 days + Prednisone (optional) 60mg each day for 5 days<br>Jaundice (pre-hepatic-post) by clinical diagnosis or suspicion of the following etiologies:<br>5.3 *Bartonella bacilliformis*: Doxycycline 100mg PO each day for 6-8 weeks<br>5.4 *Coxiella burnetti*: Doxycycline 100mg PO each day for 2 weeks<br>5.5 *Chlamydia psittaci*: Doxycycline 100mg PO each day for 7 days<br>5.6 *Salmonella typhi*: Levofloxacin 750mg PO each day for 7 days<br>5.7 *Ascaris lumbricoides*: Albendazole 400mg PO single dose (may also be related to malabsorption syndrome)<br>5.8 *Cryptosporidium parvum/hominis*: Nitazoxanide 500mg PO bid for 3 days |
|  | 5.9 *Giardia lamblia*: Tinidazole 2g PO single dose (may also be related to malabsorption syndrome)<br>5.10 *Strongyloides stercoralis*: Ivermectin 200µg/kg/day for 1-2 days<br>5.11 Trichinellosis: Albendazole 400mg bid for 8-14 days + Prednisone 60mg PO each day<br>6. Cutaneous- by papule / macule (vascular or not painful)<br>6.1 *Bartonella henselae/bacilliformis*: Doxycycline 100mg PO each day for 7 days<br>6.2 Human Papillomavirus: Podofilox gel or solution at 0.5% - apply tid for 30 days<br>6.3 *Molluscum contagiosum*: Amphotericin B 0.6mg/kg/day PO for 2 weeks OR Itraconazole 400mg/day PO for 10 weeks and then 200mg/day PO till the CD4 count in HIV patients is >100cells /µL for >6 months responding to RVT<br>By papule/macule (painful)<br>6.4 *Dermatobia hominis* / *Cordylobia anthropophaga* / *Tunga penetrans*: Petroleum jelly or an occlusive agent so when the larva moves, a manual extraction is done.<br>By papule/macule that causes itch<br>6.5 *Schistosoma* spp: Praziquantel 40mg/kg single dose PO<br>6.6 *Pediculus humanus corporis/capitis*: Ivermectin 12mg PO each day for 14 days<br>6.7 *Sarcoptes scabiei*: Permethrin cream 5% apply on skin for 8-14 hours and repeat in 1-2 weeks<br>Vesicles<br>6.8 Herpes simplex virus 1 and 2 : Acyclovir 400mg PO tid for 7-10 days<br>6.9 Sporotrichosis: Itraconazole 200mg PO each day for 2-4 weeks<br>6.10 *Rickettsia akari/prowazekii/typhi* / Leptospirosis: Doxycycline 100mg PO bid for 7 days<br>6.11 *Borrelia burgdorferi*: Doxycycline 100mg PO bid for 2 weeks<br>Morbilliform exantem<br>6.12 *Mycoplasma pneumoniae*: Doxycycline 100mg PO bid for 7 days<br>6.13 *Ehrlichia chaffeensis* / *Anaplasma phagocytophilum* 100mg PO bid for 2 weeks<br>6.14 *Histoplasma capsulatum*: Itraconazole 200mg TID for 3 days, then 200mg bid for 12 months<br>6.15 *Coccidioides posadasii*: Fluconazole 400mg PO each day for 3-12months<br>6.16 *Sporothrix* spp.: Itraconazole 200mg PO each day for 2-4 weeks<br>6.17 Actinomycetoma: Itraconazole 200mg PO each day for 2-4 weeks<br>6.18 Chromoblastomycosis (*Fonsecaea pedrosoi*): Itraconazole 300mg PO each day for 3-9 months |
| **Nutrition** |  |
|  | Mild-moderate acute malnutrition in:<br>-Children from 0 to 6 months<br>-Children from 6 months to 5 years<br>-Children 6-17 years old<br>-18 years or above: Men and women<br>-Pregnant women |

### Sample size calculations

To determine the required sample size for (1) participants aged ≥18 years, and (2) participants aged <18 years in this cross-sectional, observational study, we used 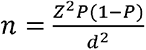 where n is the sample size, Z the confidence statistic, P is expected prevalence and d is the precision (27). Based on a past survey by the International Migration Organization, we expect 16% of migrant peoples in transit in Panama with gastrointestinal diseases; this prevalence was used for sample size calculation (28). The sampling frame composed of a total population expected to enter the MRS (2500; 20% of those are <18 years). In total, 120 participants <18 years and 160 participants ≥18 years will be included. We have a larger planned sample for the questionnaire, as we expect over half of these individuals will consent to filling out the questionnaire but not be willing to participate in the more time-consuming clinical components.

### Ethical considerations

This study has approval from the *Comité de Bioética de la Investigación del Instituto Conmemorativo Gorgas de Estudios de la Salud* (260/CBI/ICGES/21). Participants 18 years and older will be asked to read a study information sheet and sign the informed consent form. Participants 12-17 years will have their guardian read and sign the guardian information sheet and informed consent form, and the participant will be required to read and sign the informed assent form. Participants <12 years will be required to have a guardian read and sign the informed guardian consent form. Participants will not be offered monetary compensation for their participation. However, a small snack (valued at less than $3.00) will be provided. All participants will be given educational sheets that cover the syndromes and infections tested for, and these sheets will be posted publicly at the MRSs. The consent forms, assent forms, questionnaire, and educational materials will be available in Spanish, English, French, Haitian Creole, and Portuguese languages.

### Participant selection

In Component A, we will use purposive sampling. Community leaders will be asked to attend *focus group interviews*, and institutional, national, and organization leaders will share their perspectives through *individual semi-structured interviews*. A group of migrant peoples will also be selected to participate in *individual semi-structured interviews*. Select migrants accessing health services may be accompanied to observe their experiences within available healthcare providers. A total of 70 participant interviews and focus group discussions are anticipated within the qualitative study component, as this number is expected to allow us to reach saturation in capturing stakeholder perspectives and experiences.

In Component B, participants will be included systematically, where groups of 40-50 individuals arrive at the MRS where the sampling will be undertaken. When lined up to be registered with the Migration Service, small invitation cards will be given to every 1:*n* person from each stratum. The *n* will be calculated at the start of each day and depends on the number of expected strata to arrive each day, to include between 30-50 participants daily. After the person finalizes the migration registration process, they will be asked to join the study group at the respective study tents. Inclusion and exclusion criteria are dependent on the study component, as outlined in Table 2. As potential participants may be at the MRS for an undetermined amount of time, this study will only include participants who do not have a planned exit from the MRS within 8 hours.

### Data capture

Data capture will differ by study component and methods employed. Component A will include participant observation, semi-structured interviews, and focus groups captured via ethnographic fieldnotes and recorded on a digital recorder. Interviews and focus groups will be conducted in Spanish and, if needed, interviews with migrant persons will be undertaken in English and through real-time interpreters of French, Portuguese, or Haitian Creole. Component B will include a self-administered questionnaire on a tablet using *Kobo Toolbox Software* (Harvard Humanitarian Initiative, Cambridge, MA), and will be stored on the device under the participant’s code until wifi is available. Clinical and laboratory information will be inputed into *Kobo Toolbox* with a separate entry for each participant.

### Pharmacological and psychological treatment and referral

On-site treatment by study subcomponent is found in Table 5. If the participant needs to be referred to primary, secondary or teritiary facilities for further care (Supplementary Table B), this will be organized by the study team and the participant will be transported with no cost to them.

#### Pharmacological treatment

On-site pharmacological treatment will be given according to the severity of the affectation. A higher level of care will be arranged if needed to on-site or referral to the nearest health center in the town of Meteti or to the secondary and tertiary hospitals, in Chepo and Panama City, respectively. The field clinic where the study is undertaken is rudimentary and will not attend to all needs. Therefore some cases will be referred to health centers and hospitals where a) the condition requires further testing or specialized care, or b) treatment is intravenous or daily injections, as described in Table 5.

#### Psychiatric care and treatment

Following the RHS-15 screening and meeting with the psychiatrist, individuals who would like to participate in group-oriented therapy are welcome to attend language-based sessions. Additionally, at any time, if the participant seems in distress, presents with acute psychosis or self/heteroagression, immediate consultation with an on-site psychiatrist will be done. Treatment protocol is summarized in Table 5 according to syndromic presentations.

#### On-site management of distress

At any time during the study, if a participant is anxious, depressed, or distressed, the “Distress Protocol” will be activated (Supplementary material C). The “Violence Protocol” will be implemented for individuals who report sexual violence during the qualitative component, questionnaire, or clinical exam (Supplementary material D). These are based on the Ministry of Health procedure for sexual violence, rape, incest, and a previously published protocol for children who report violence that has been used in previous studies in Panama and worldwide (23, 29).

#### Lab and clinical results

The laboratory POC tests and clinical results will be given to every participant in paper format the day they are included in the studies. All results will be explained to the participants at the time results are offered. The results will also be found in a password-protected location on the Gorgas Memorial Institute for Health Studies’ webpage. Each participant will have access to a secure portal for one year, which will house their study results and educational sheets. The portal will be accessible from a phone or computer. The participants’ access information and password will be clipped into their passports.

#### Data analysis plan

Data analysis will depend on the study component.

For Component A, recordings and fieldnotes will be transcribed; all identifying information will be deleted. The recorded files will be destroyed after the transcription. Transcripts will be coded by MB and AG using *Nvivo* software. Both coders will work independently and codes will be compared, discussed and discrepancies will be resolved. Transcripts will be saved on an encrypted external hard drive. The data will be analyzed with a thematic analysis method and in an essentialist manner, meaning the participant experiences will be revealed. Transcripts will then be coded at a manifestation level to include the main themes. It is also possible that some codes emerge inductively from the data. This hybrid method of analysis from the data has been previously described and demonstrated to be rigorous (30). A thematic map will be constructed, revised, and altered based on team discussions; contradictions and negative cases will be identified and presented.

For Component B, descriptive and analytical methods will be used to describe the findings of the quantitative questionnaire, including sociodemographic factors, sexual and reproductive biography, sexual behaviors, non-consensual sexual relations, self-reported health, family whereabouts, and social wellbeing. In addition, the prevalence of laboratory and clinical diagnoses will be presented for the sample as a whole, as well as for key subgroups, including men, women, adults and young persons. Quantitative questionnaire responses will be used to describe the epidemiology of test positivity and diagnoses. Sociodemographic and behavioral variables that are significantly associated with health outcomes of interest in bivariable analyses will be included in multivariable regression models to estimate strength of adjusted associations and test their significance.

## Discussion

By 2050, the UN estimates that over 1 billion people could be displaced due to climate change alone (31, 32). Many others are leaving their country of origin for work, family, conflict, persecution, and natural disasters (32). This increase in population movement is already evident throughout the Americas, especially in Panama, where migrants from different Caribbean regions, South America, Africa, and Asia, travel through South and Central America en route to North America. Unfortunately, the world and Latin America are not prepared to provide medical care for mass migration.

PAHO has highlighted a need to strengthen both, health surveillance and the monitoring of existing programs targeted towards migrant peoples in the region (16). This rapid evaluation of health status and service provision among migrant peoples in transit aims to respond to this evidence gap. Employing a mixed qualitative and quantative approach, the study will investigate the health status and needs among migrant peoples in transit passing through the Darien MRSs. Currently, individuals with acute illness or emergency health conditions, such as pregnancy, may be transferred to nearby MINSA health centers. However, the occurrence of other health conditions, such as anxiety, PTSD, depression, genito-urinary syndromes and tropical diseases, as well as mal/under-nutrition, remain largely undetected and ignored; information related to their prevalence among migrant peoples in transit through the region remain widely unknown.

As the demographics of migrants who opt for this perilous route shift, information related to the health needs of this population and opportunities for strengthening service provision at the MRS and regionally for migrant peoples in transit is essential. This study is critical to understand health priorities, especially with infectious and tropical disease, nutrition, mental health and sexual and reproductive health of those who pass thorough the DF. Additionally, findings could be used to optimize scarce resources targeted towards improving the wellbeing of migrant peoples who pass through this region. Previous research among people in transit in Central America and Mexico has led to recommendations to decriminalize irregular migration (33); identify and stop human trafficking, particularly of women and children (33); and expand the number of migrants’ shelters as a way to meet basic needs (34). Further, studies recommend better coordination and collaboration among governmental and non-governmental organizations—and across borders—to prevent and control health problems faced by migrant people in transit (34). Further, researchers recommend improving access to primary healthcare, including STI and HIV testing, among this population (34, 35). To ensure that the results of this project inform programs and policies for migrants in transit, results and targeted intervention recommendations that will have emerged from the qualitative and quantitative studies will be presented locally to MINSA and international non-governmental organizations that focu on service provision to migrant peoples.

Access to health care is a human right—increasing migrant health and health literacy translate into future social engagement, citizenship, and productivity (36). The overarching goal of this project is to document the health needs of migrant peoples in transit who pass through Panama in order to provide initial surveillance information and recommendations for targeted interventions for this mobile population. To do this, the project will use point of care testing and a comprehensive, easily deployable package of health services. Strengthening point of care diagnosis and service delivery will be essential to address barriers to quality health care for migrant peoples that frequently have insufficient time to receive provider follow-up and continuity of care. The literature from other contexts of migration indicates that strategies to screen and treat migrant populations for transmissible diseases, including tuberculosis, hepatitis B, hepatitis C, HIV infection, enteric bacteria, and helminths, can be both impactful and cost-effective (37) in the control of infectious disease. Addressing these unmet health needs and improving service provision can help alleviate the incredibly marked inequities migrant peoples have experienced and sustained in their journey.

The challenges to deploy and deliver a point of health care for migrant peoples in transit are enormous—facing barriers including discrimination, violence, communication and insufficient access to healthcare (4, 38). Such issues have described a perceived idea of ‘deservingness’ that categorizes patients, particularly migrant peoples, as more or less deserving of access to quality health care (39). There are few published data on healthcare needs of migrant peoples in transit through Latin America in route to USA. Grey literature from UN and other international NGOs suggest the greatest health needs include treatment of fever, skin problems, diarrhea and prenatal care. The successful implementation of a health screening protocol like our proposed study may become a platform to scale up in different migratory routes in the Americas and in other regions. Furthemore, the successful conduction of this study may provide crucial information to increase awareness and lead to effective targeted healthcare services for migrant peoples in transit.

## Conclusion

The results of the qualitative and quantitative studies answer the PAHO call to contribute to a better understanding of the health status and health needs of migrant peoples in transit through Panama. It will also serve to strengthen interventions of national and decentralized health and surveillance systems nationally and regionally. Most importatly, the results from this study can be used to develop targeted interventions of healthcare provision for migrant peoples in transit through the DF and the Americas.

## Data Availability

No data has been collected yet.

## Acknowledgments

We are thankful to the Center for Research and Diagnosis of Emerging and Infectious Diseases, Meteti, Darien of the *Instituto Conmemorativo Gorgas de Estudios de la Salud*, Community Development Network of the Americas, *UNICEF, Médecins Sans Frontières, Panamás Ministerio de Salud, Servicio Nacional de Fronteras* (SENAFRONT) and *Servicio Nacional de Migración* for their ongoing support. We are additionally thankful to those who have contributed thus far to this project: Acino Swiss Farma, Ann Knudson, and the Oklahoma University’s Physicians Pharmacy, Sagrav SA, and Omega Diagnostics.

## Declaration of conflicting interests

The authors declare no conflicts of interest.

## Funding

Funding is provided by *Instituto Conmemorativo Gorgas de Estudios de la Salud* (Panama), Community Development Network of the Americas, Acino Swiss Farma (donation), Neglected Tropical Diseases Fund and University of Colorado (donation by Mr. Howard Janzen). Additional donations have come from Sagrav SA and Omega Diagnostics, and a large proportion of the diagnostic tests have been donated through individual research funds from the National Research System (’SNÍ by member Dr. José Antonio Suárez).

## Author contributions

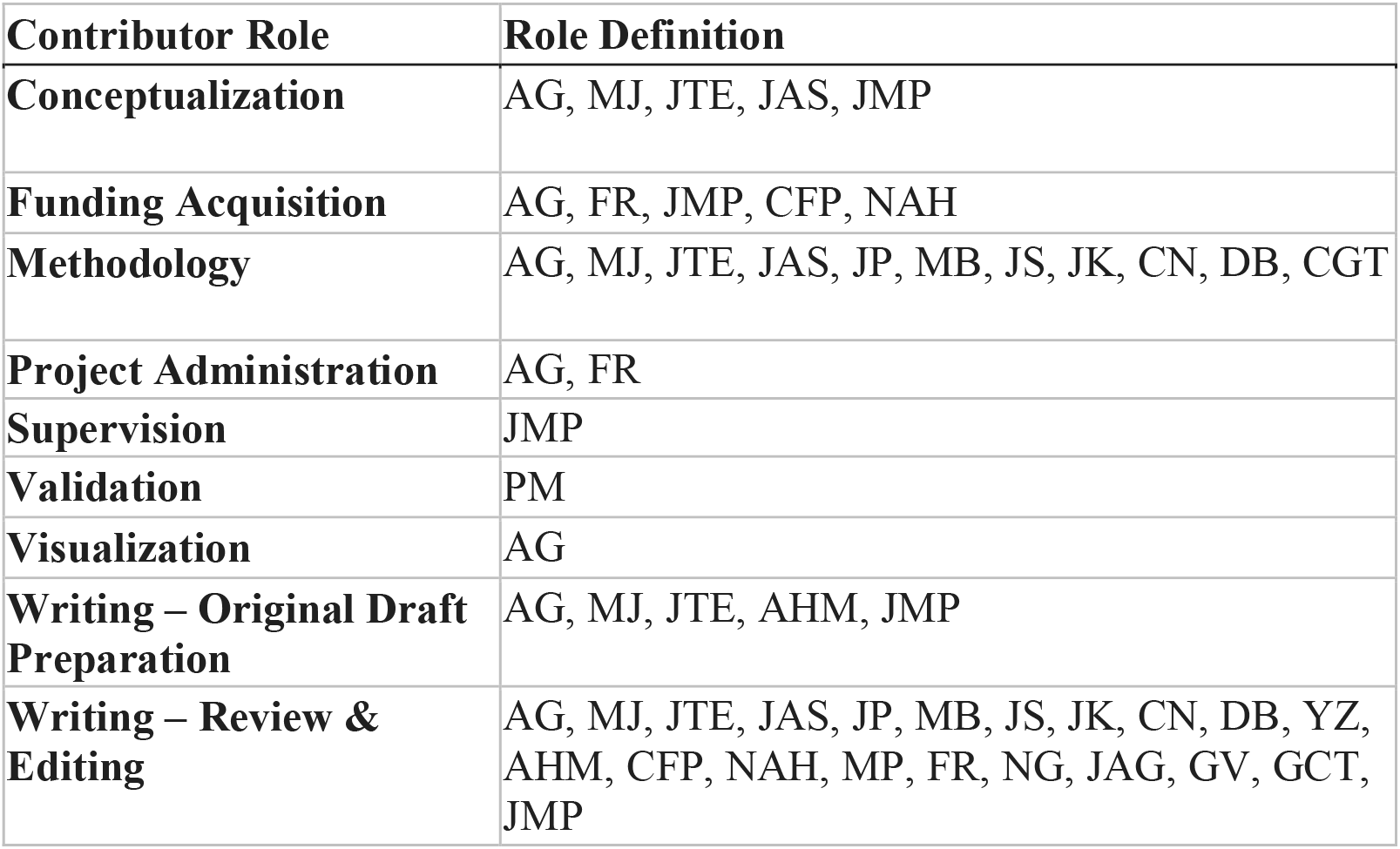

---

**Map 1:**
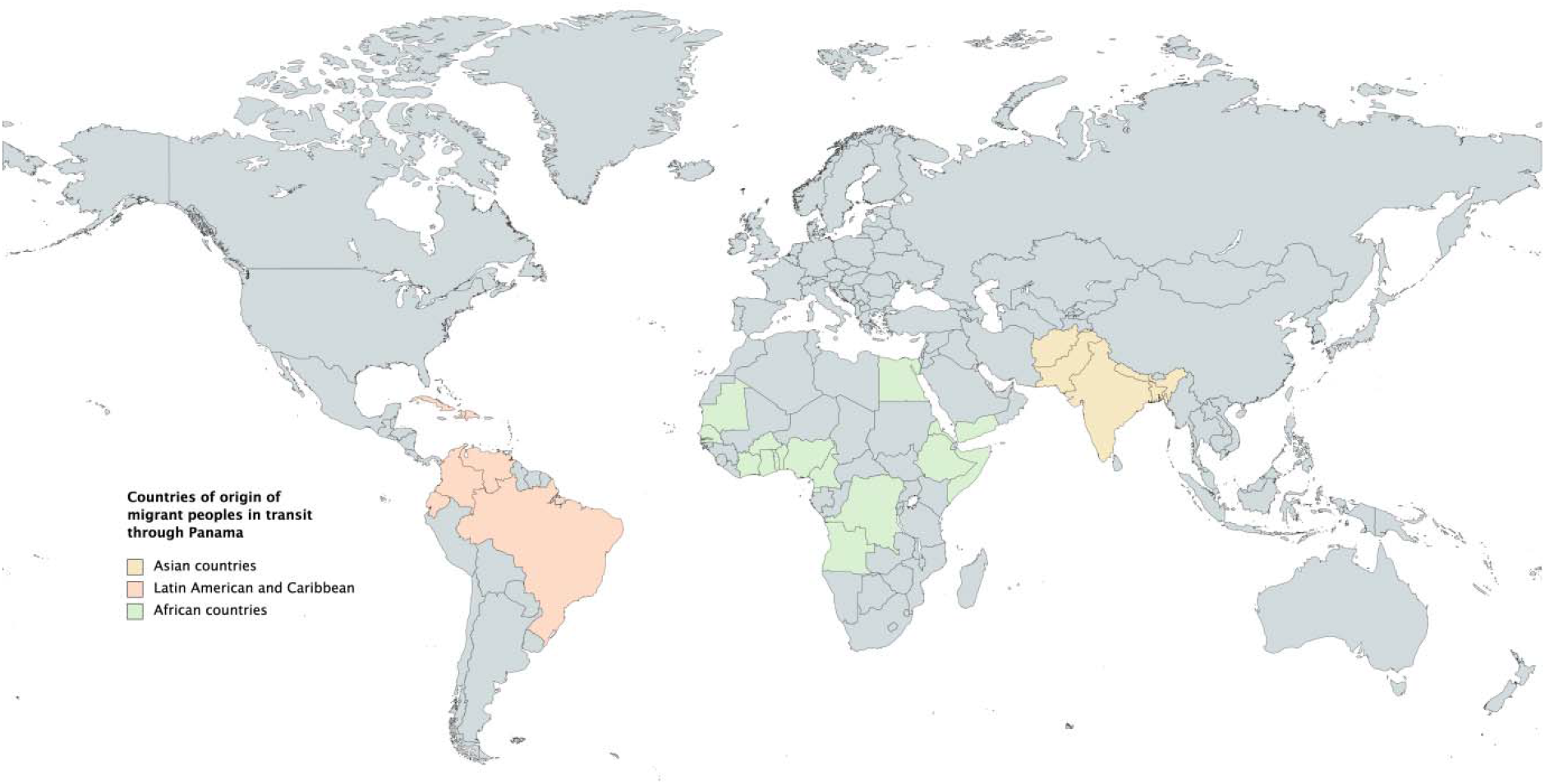
Countries of origin of migrant peoples in transit, after crossing the Darien Forest, Migrant Reception Stations in Darien, Panama (documented from data collected by Migration System, Panama, March 2021).

**Supplementary material A:** Self applied questionnaire (English version, however questionnaire will also be available in Spanish, Haitian Creole, French and Portuguese). Note: this questionnaire will be self-applied on a tablet, therefore the format will differ and conditioned questions will only only appear if they apply. This questionnaire was developed from existing surveys and modified for migrant peoples in transit through the Darien Forest, Panama (17–26).

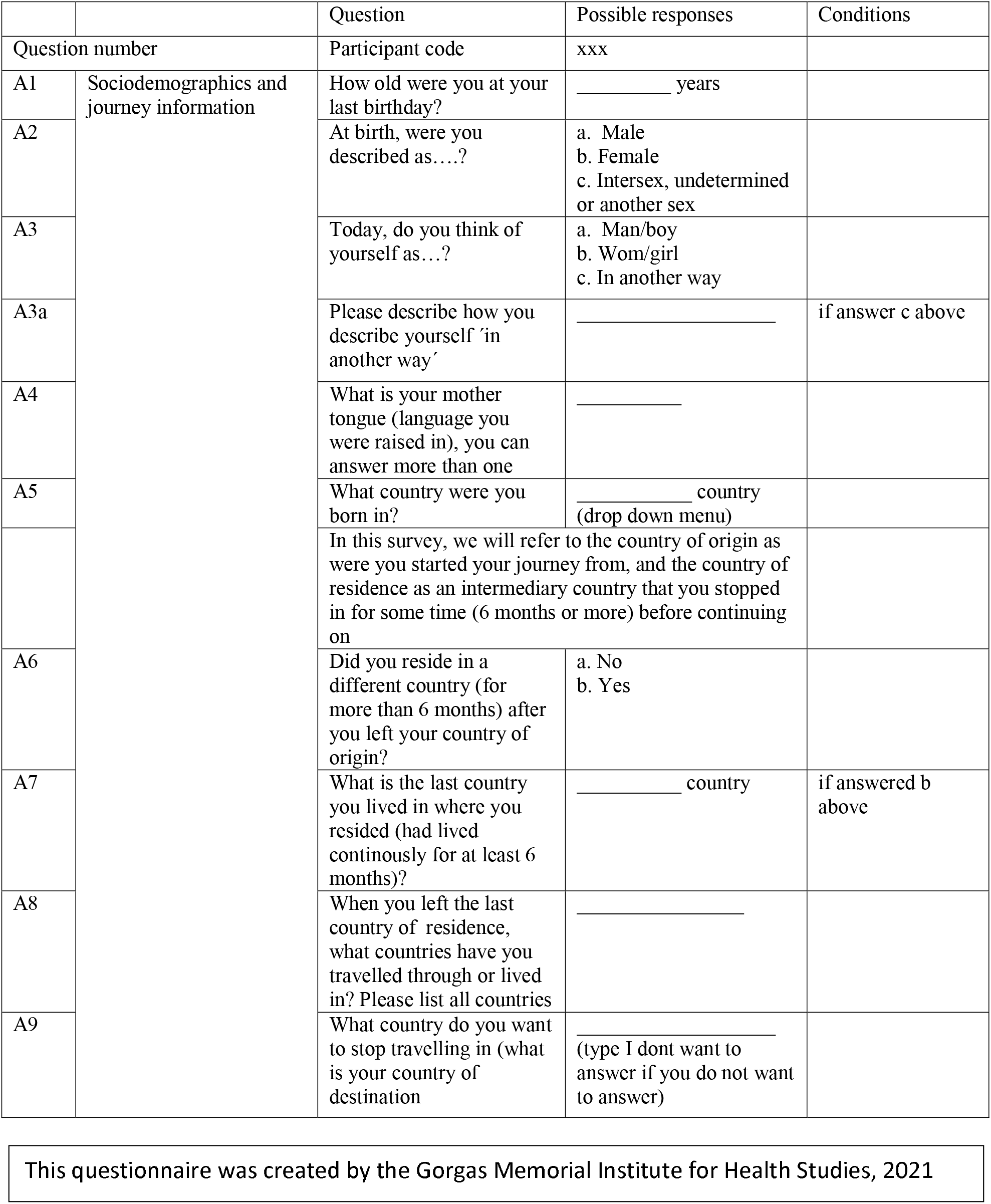
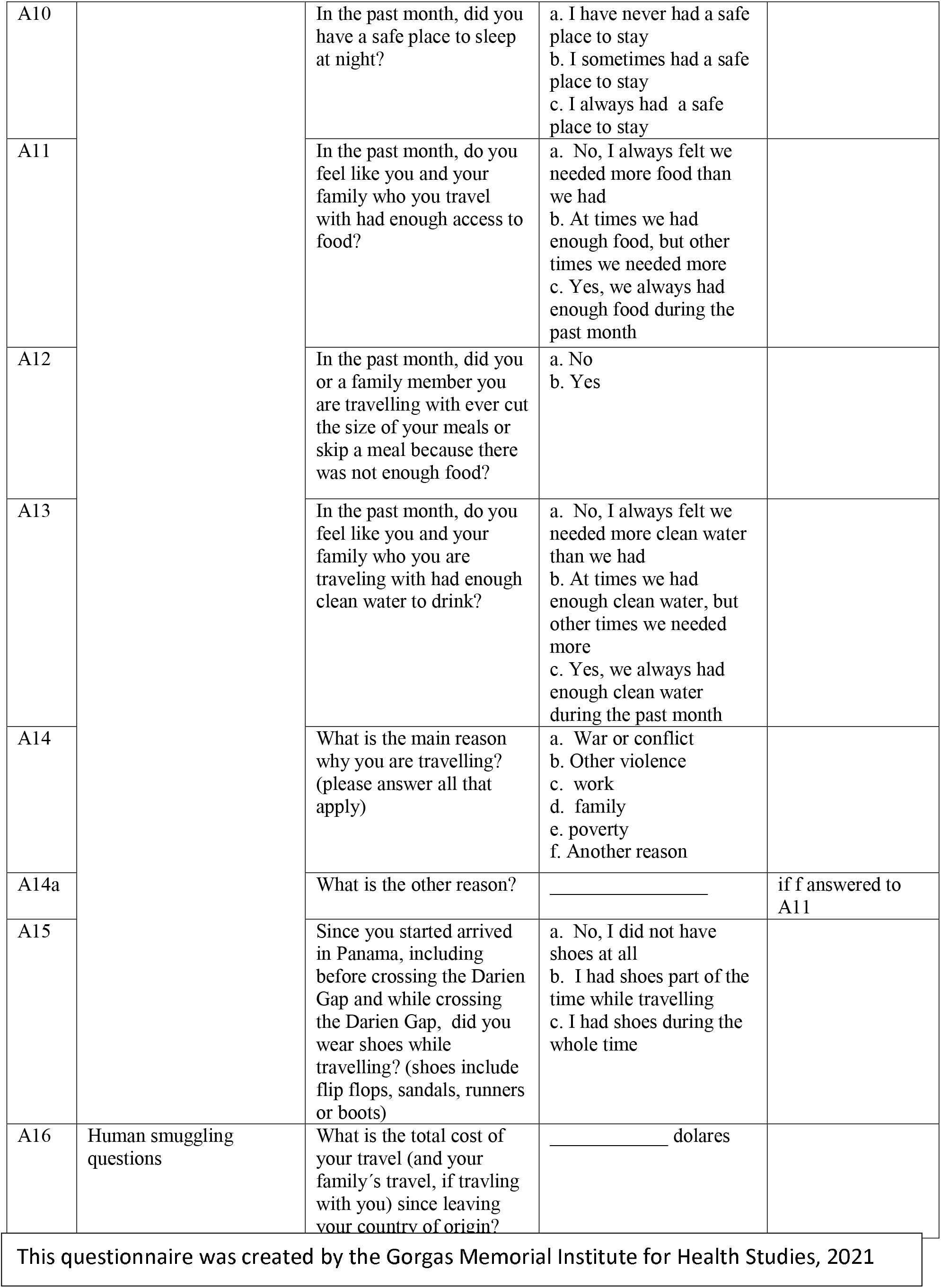
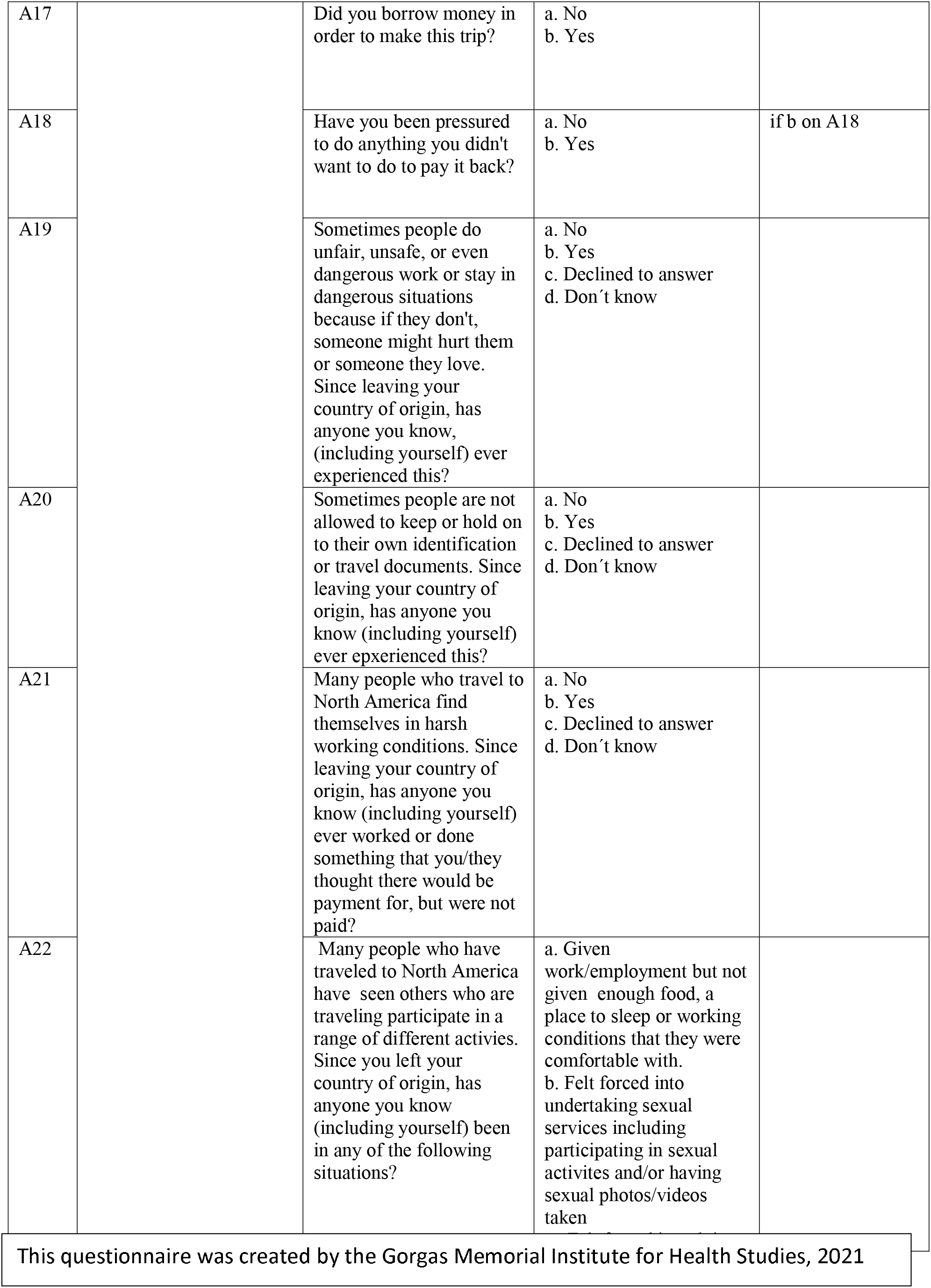
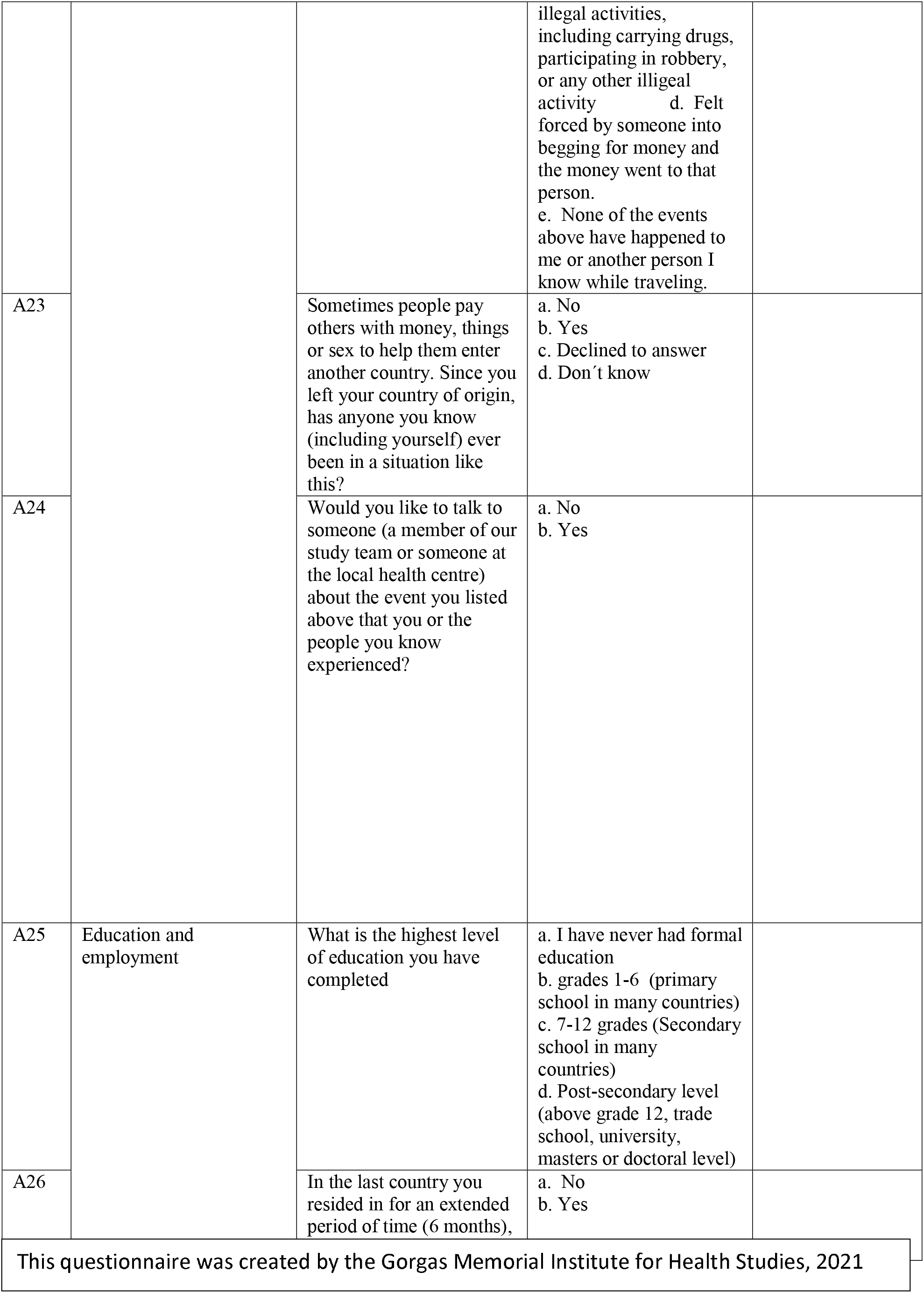
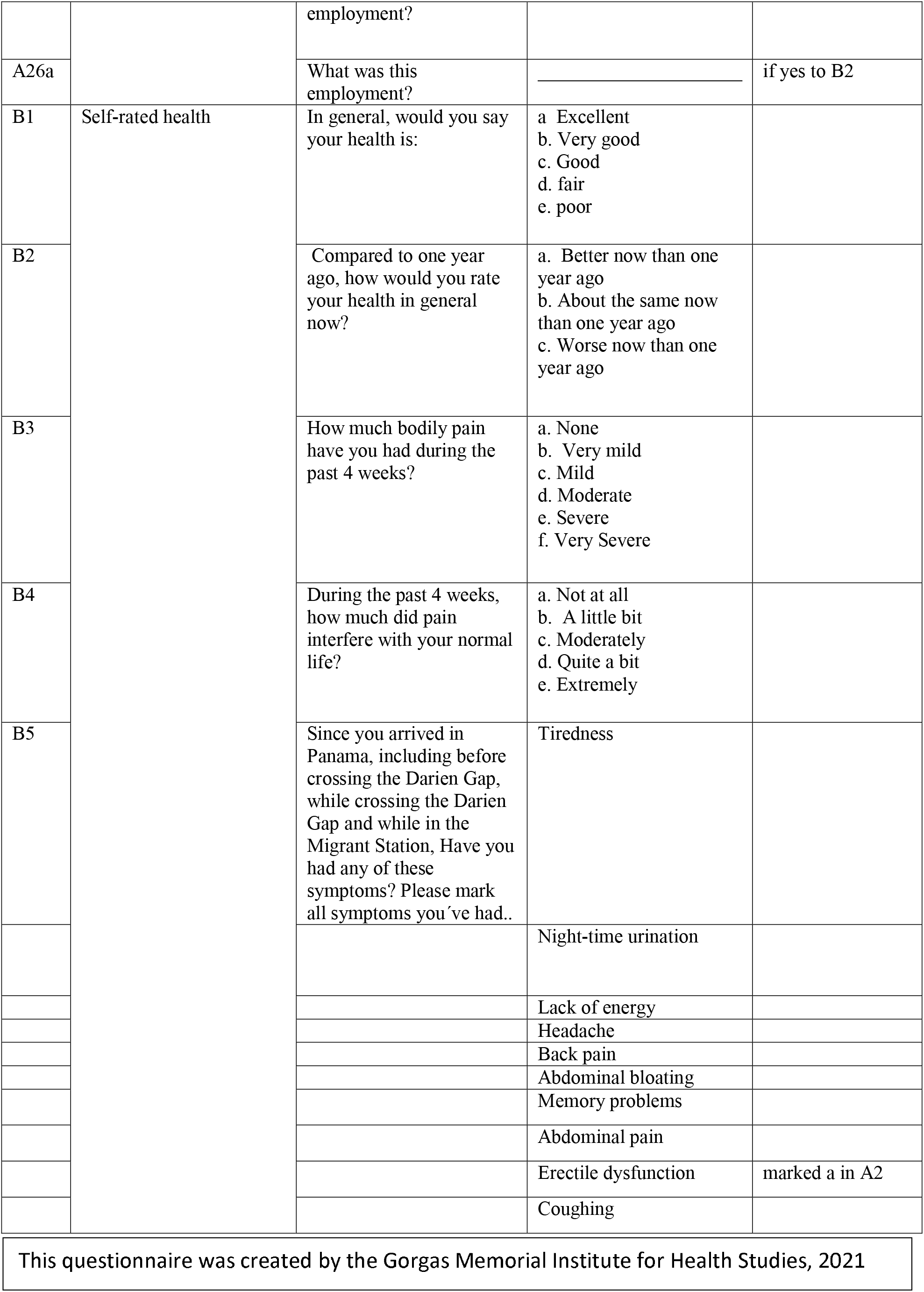
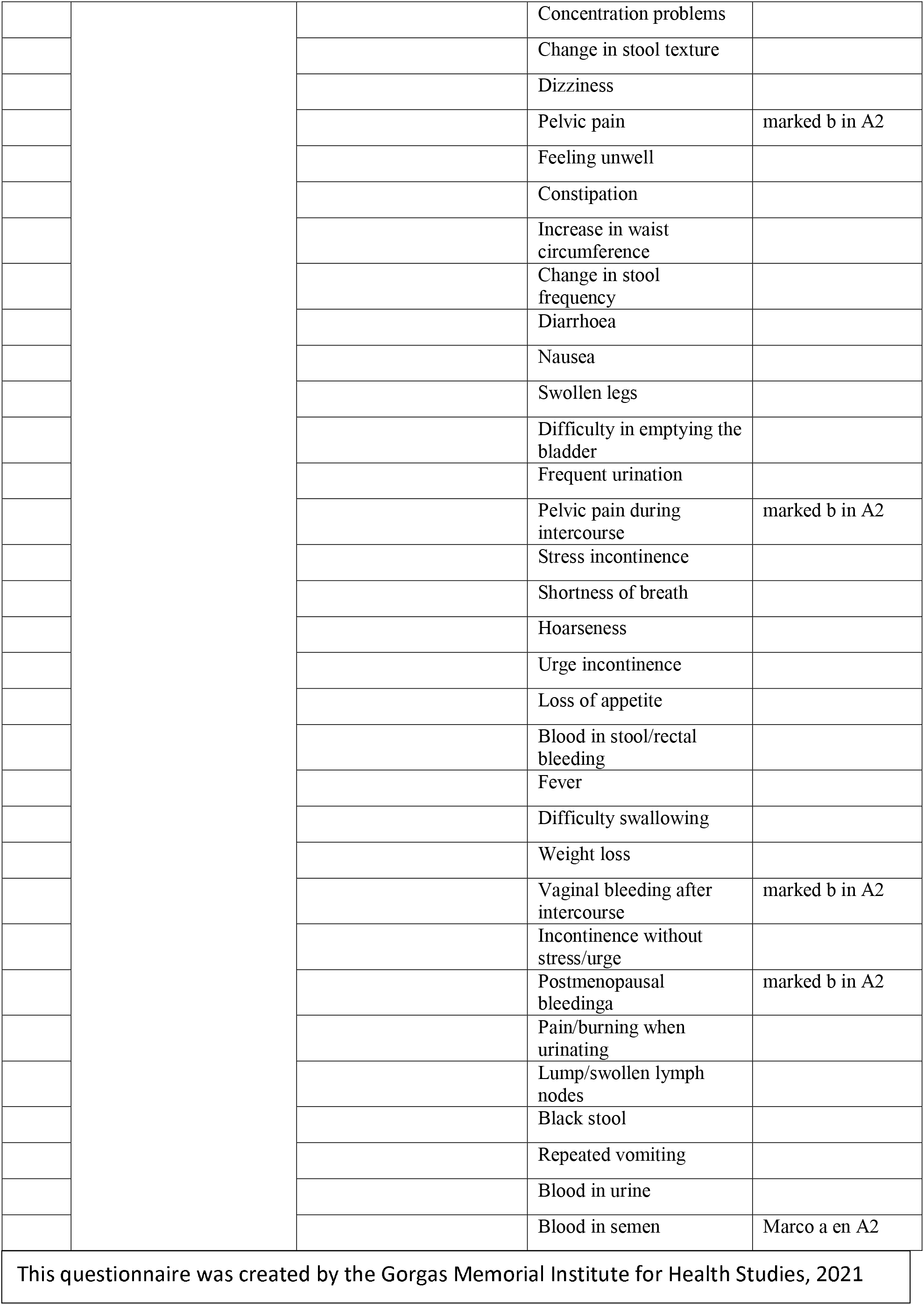
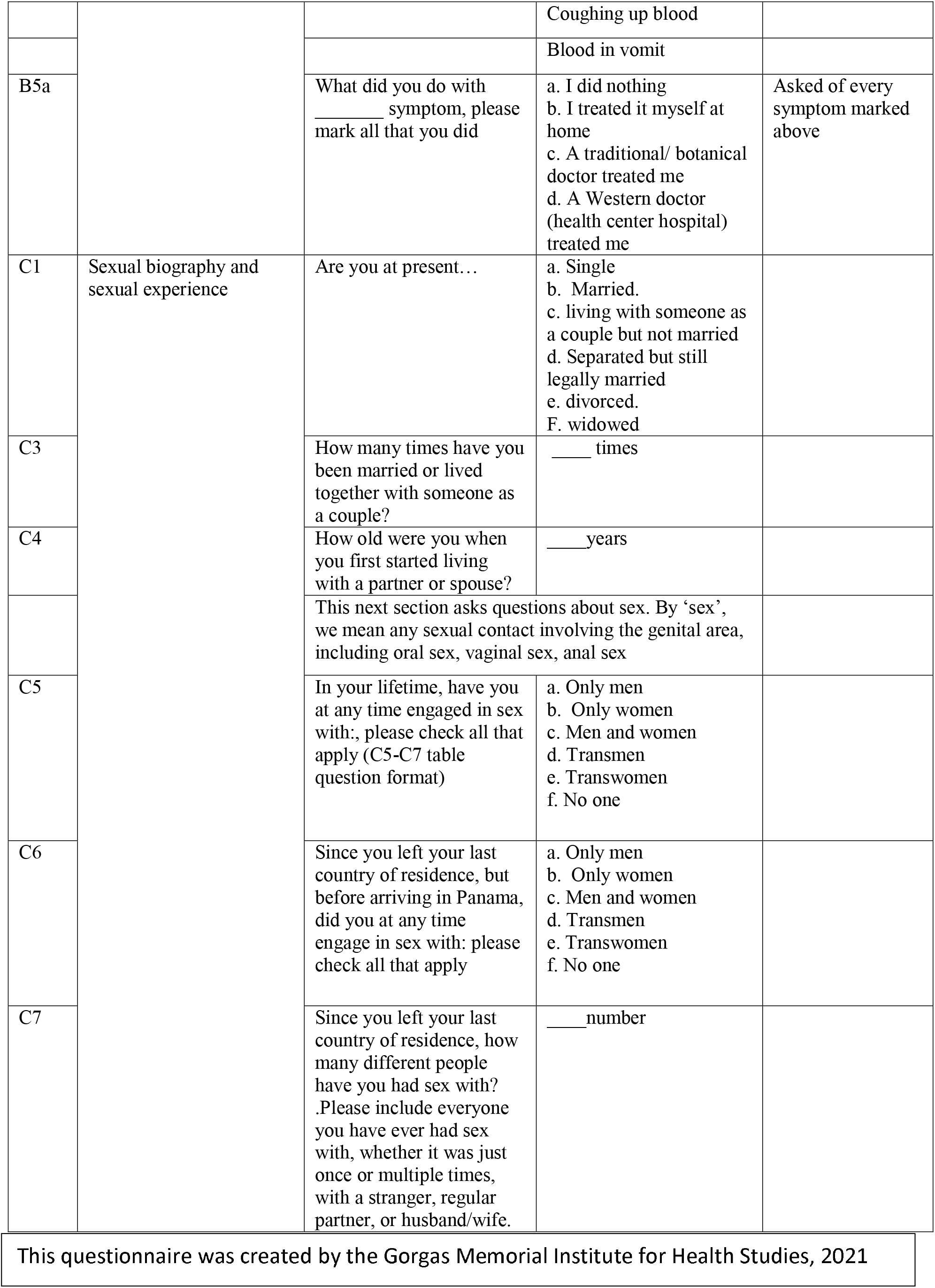
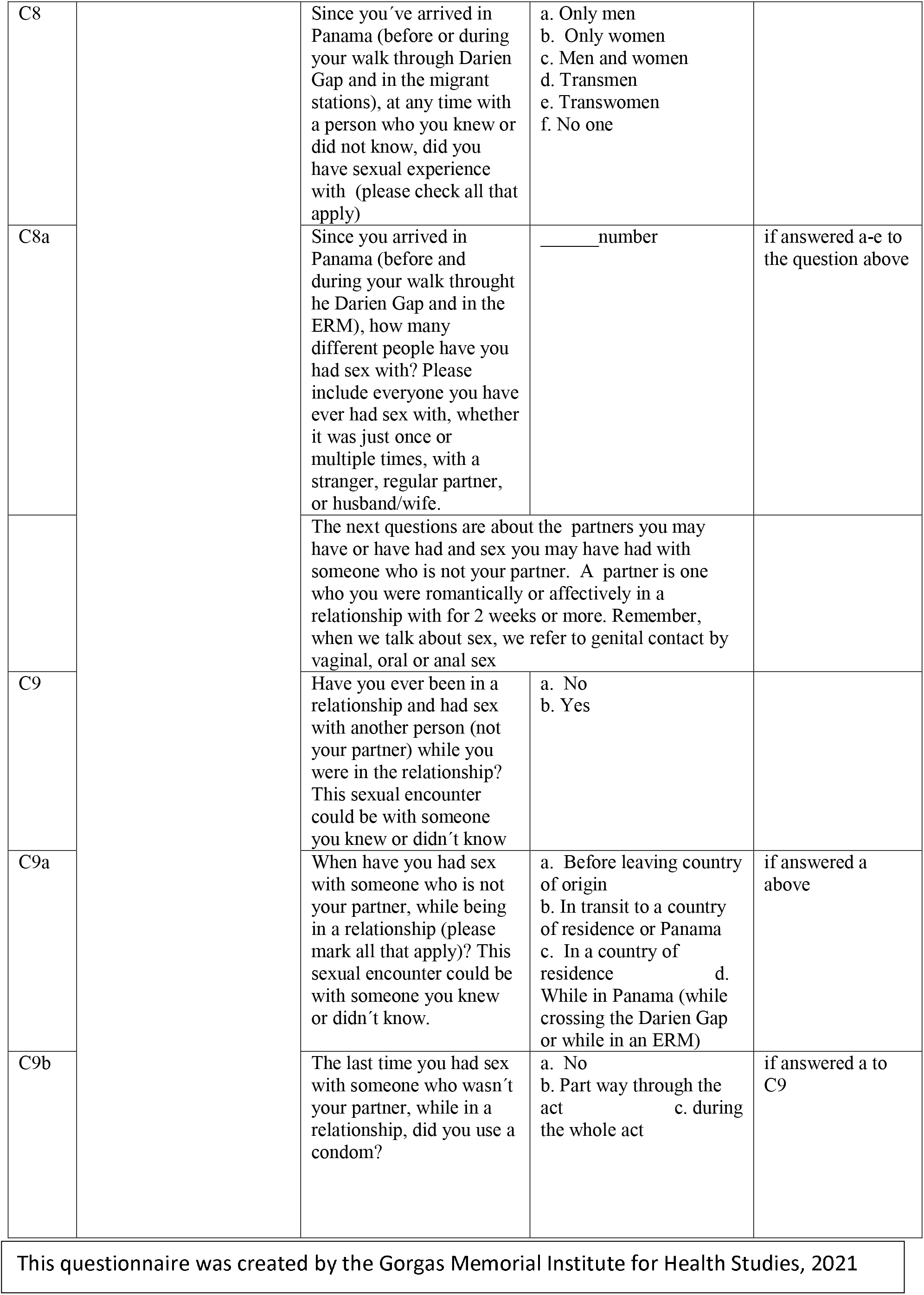
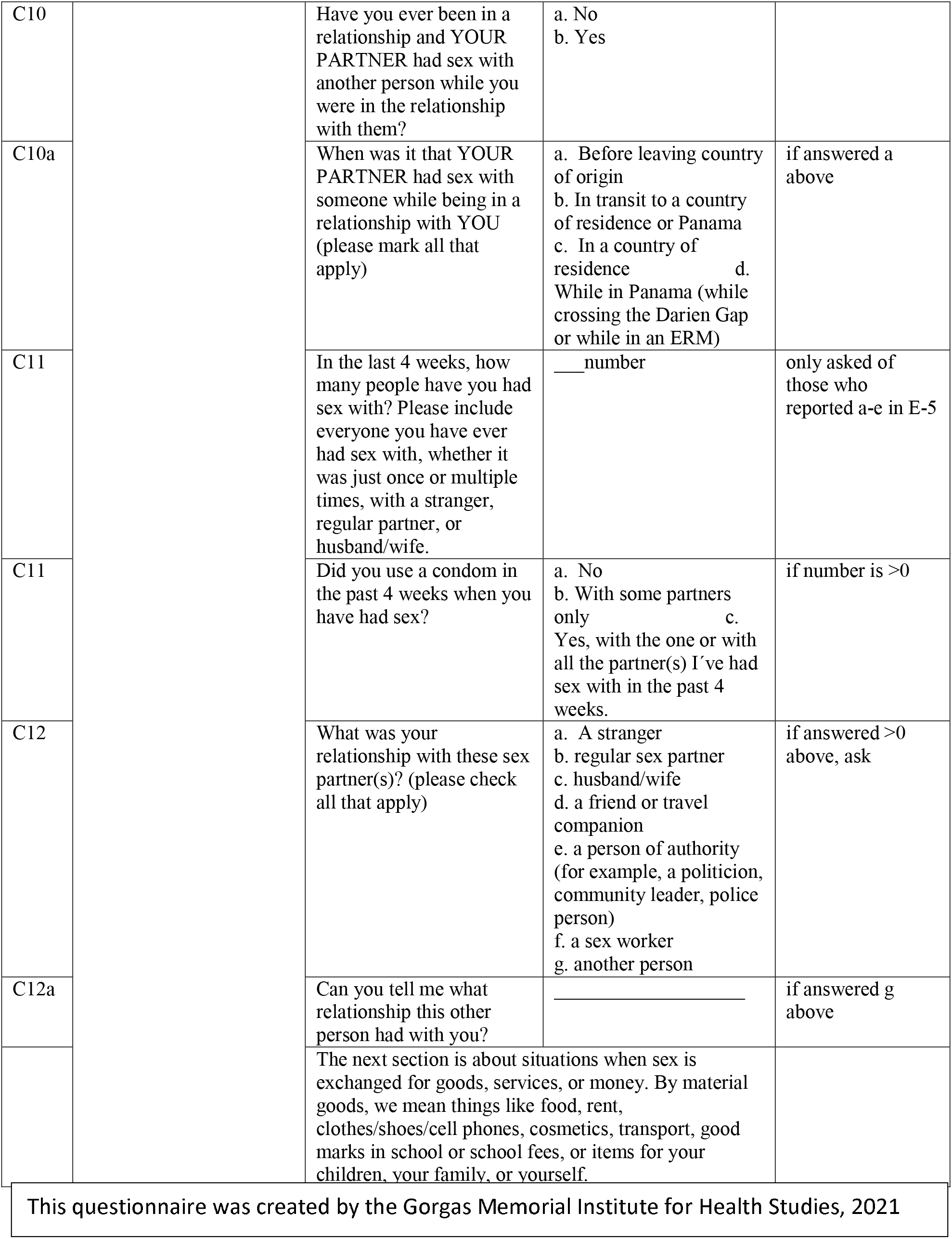
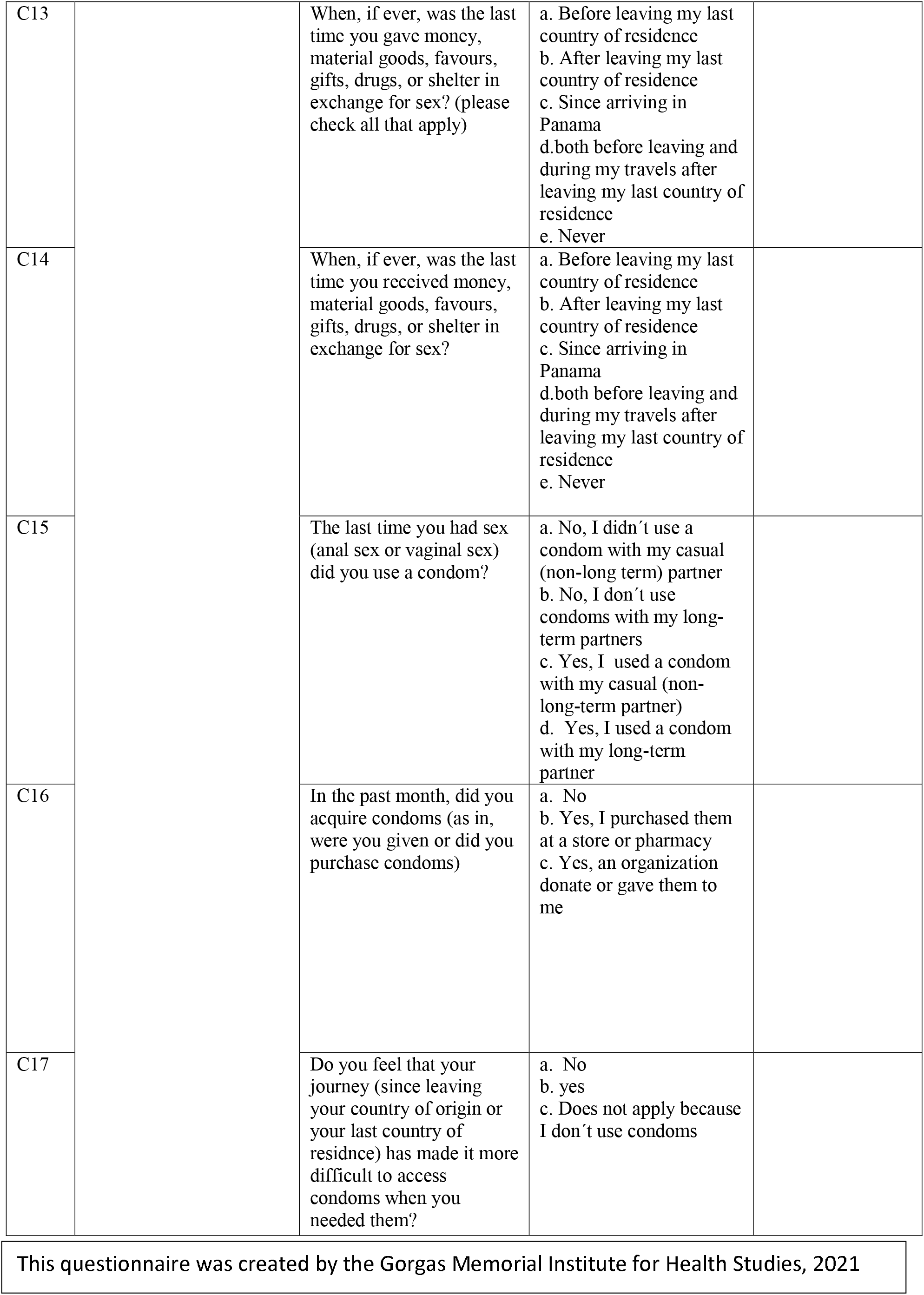
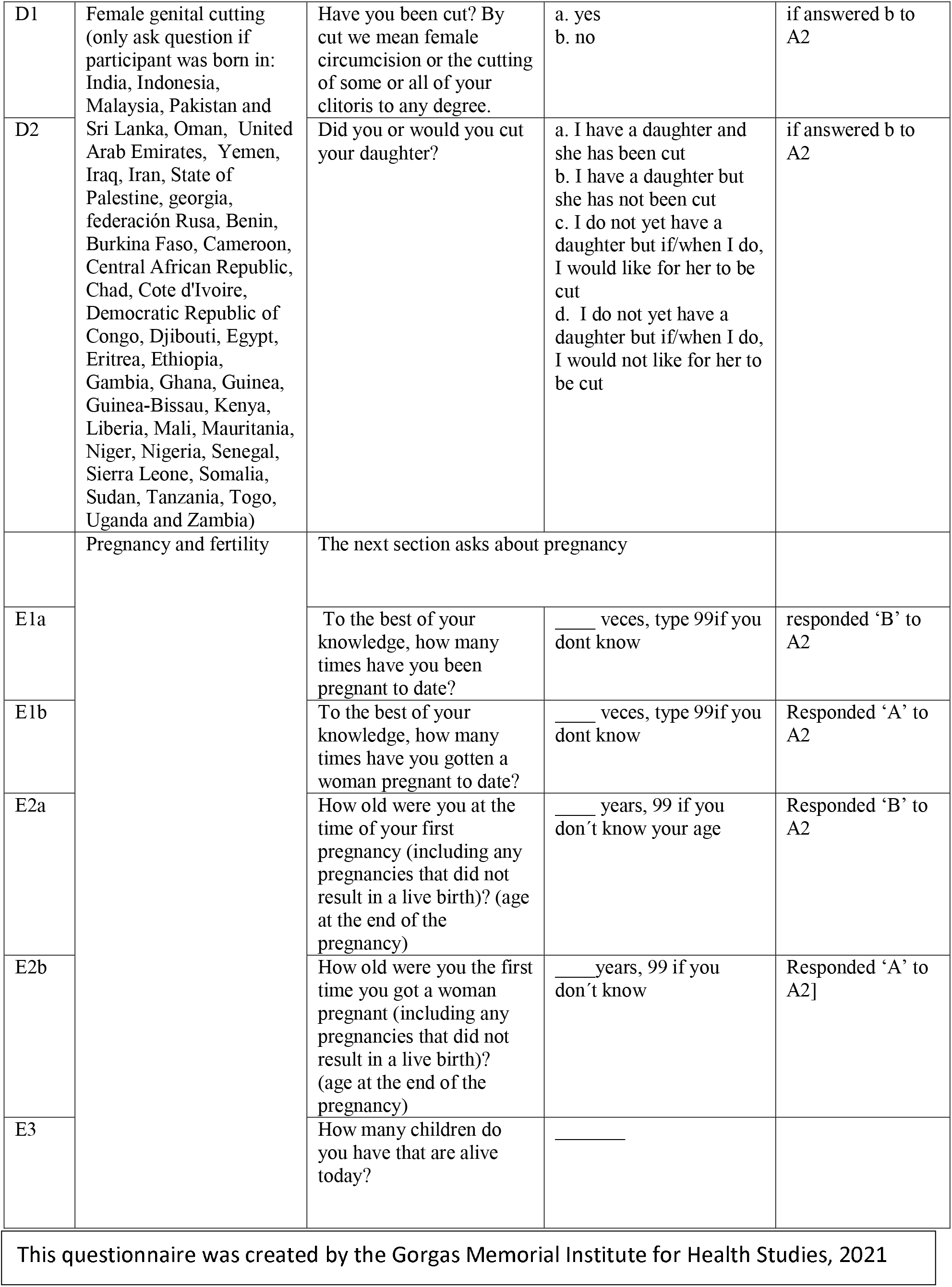
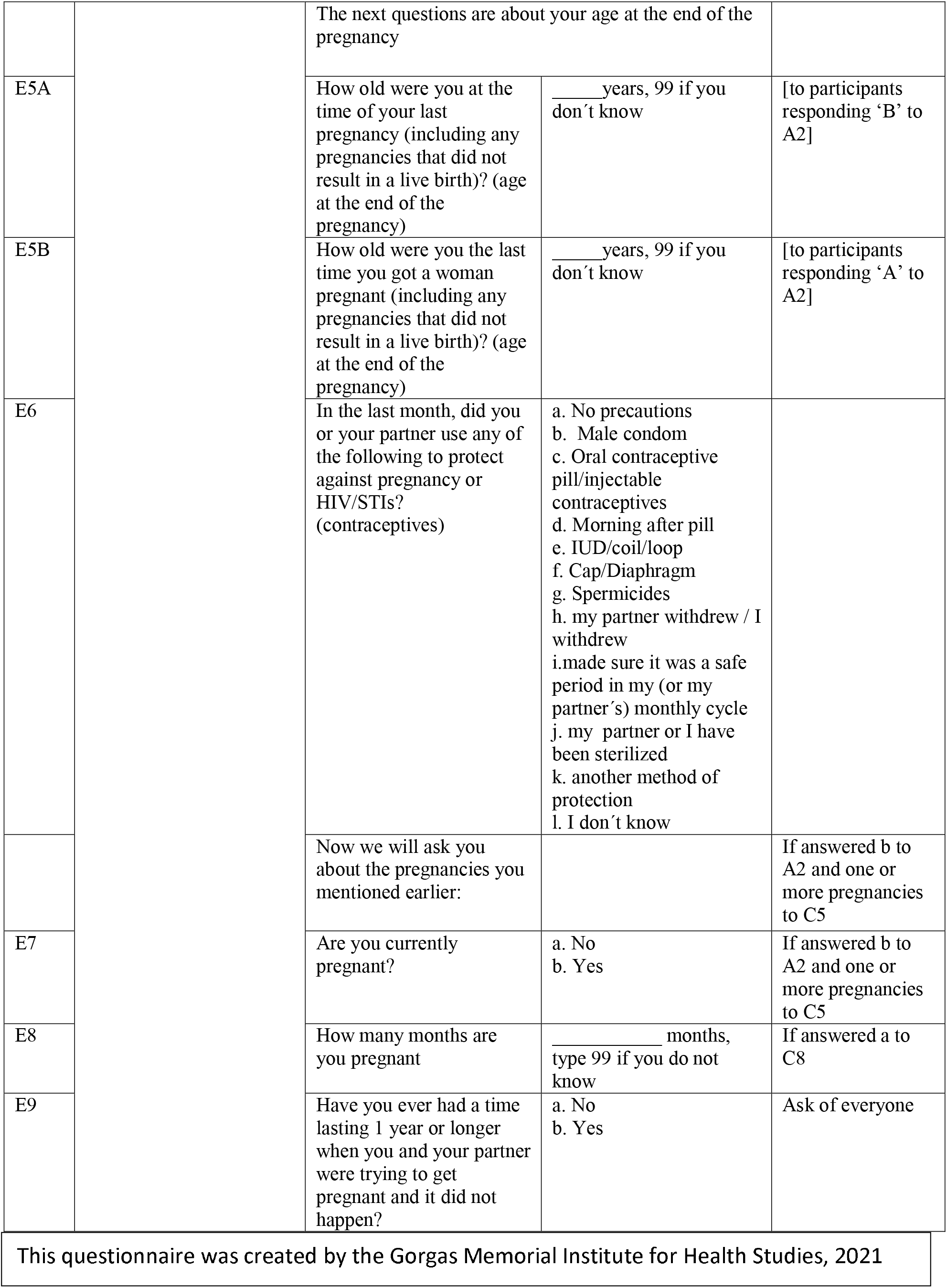
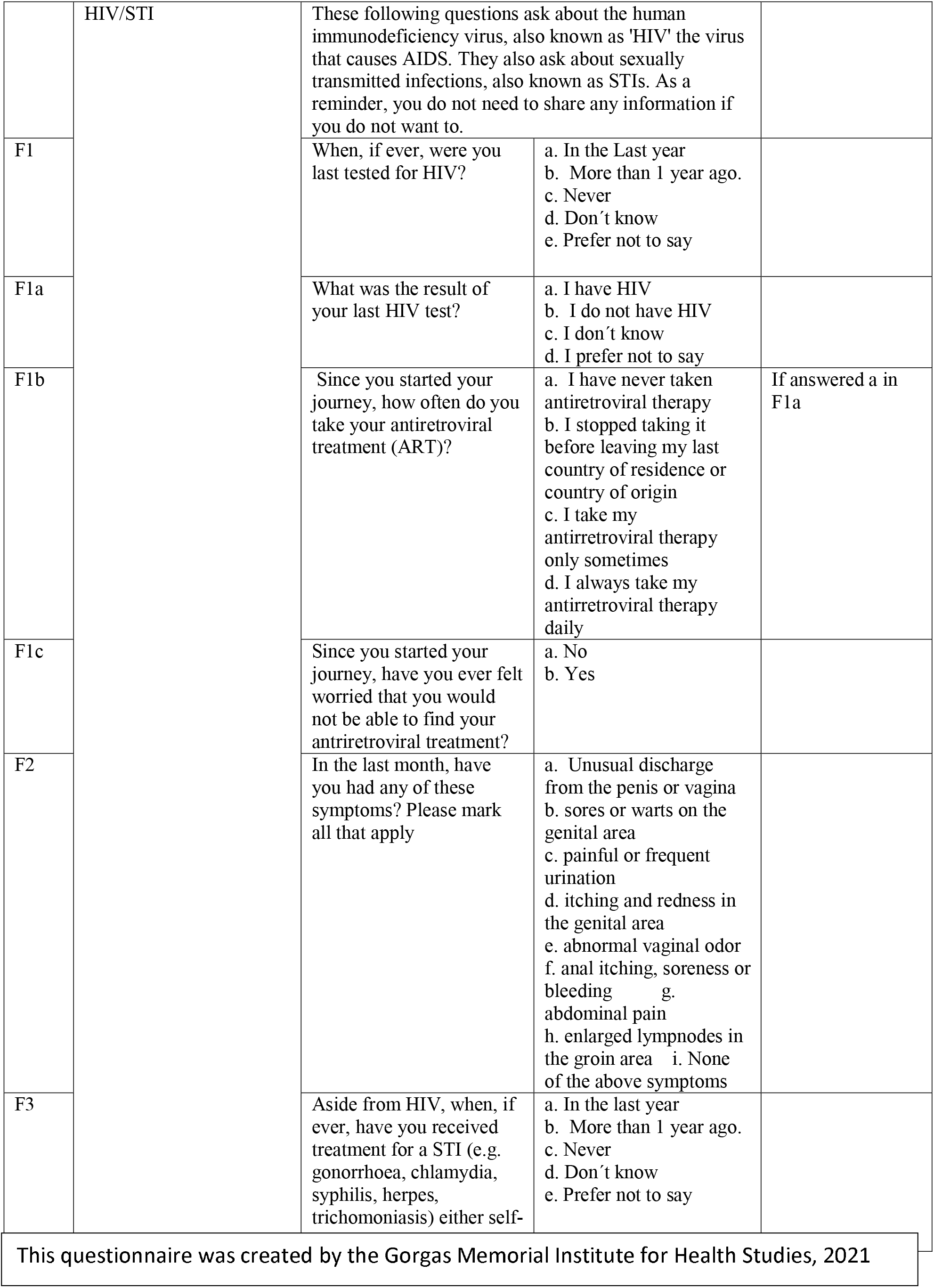
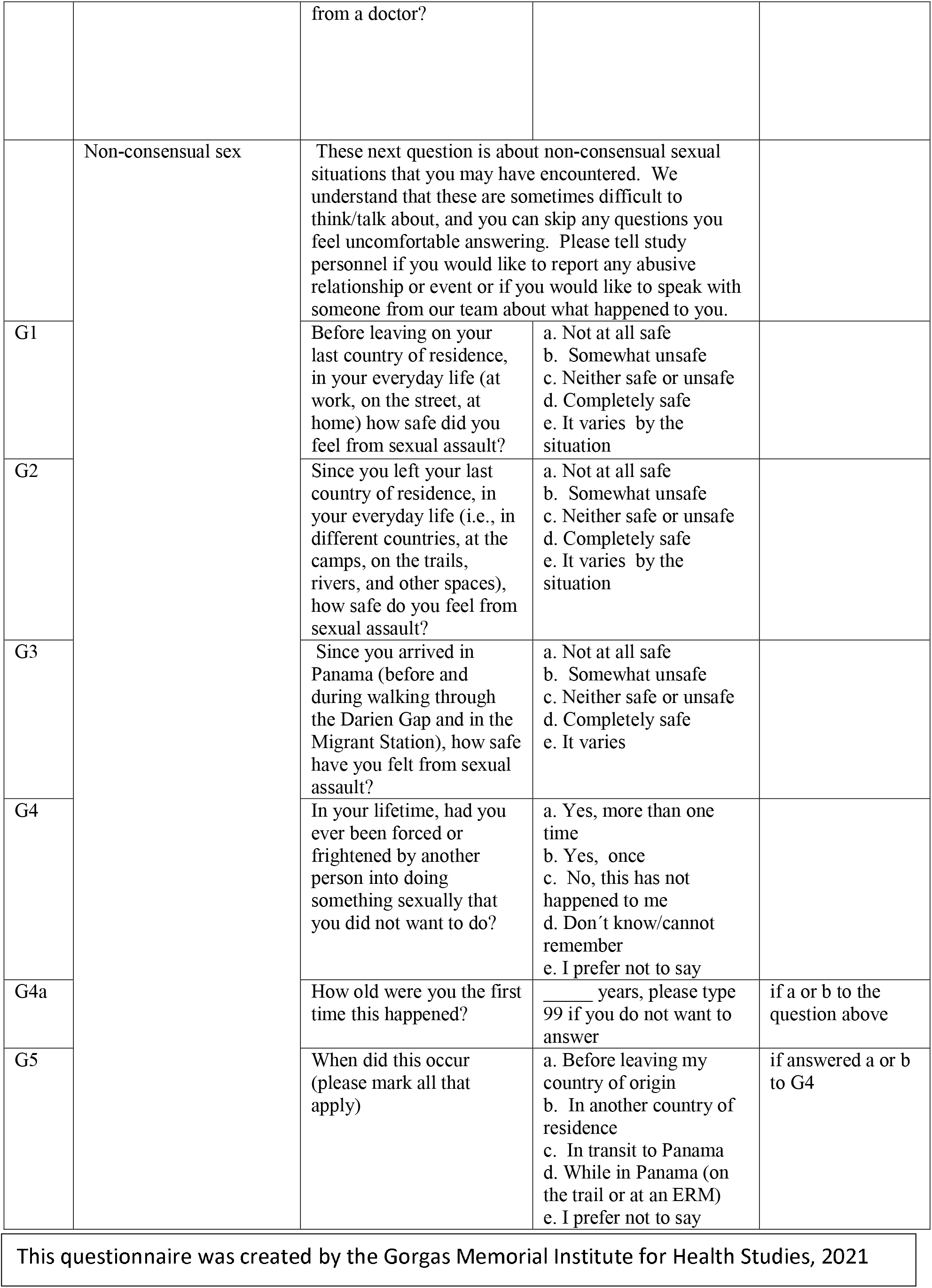
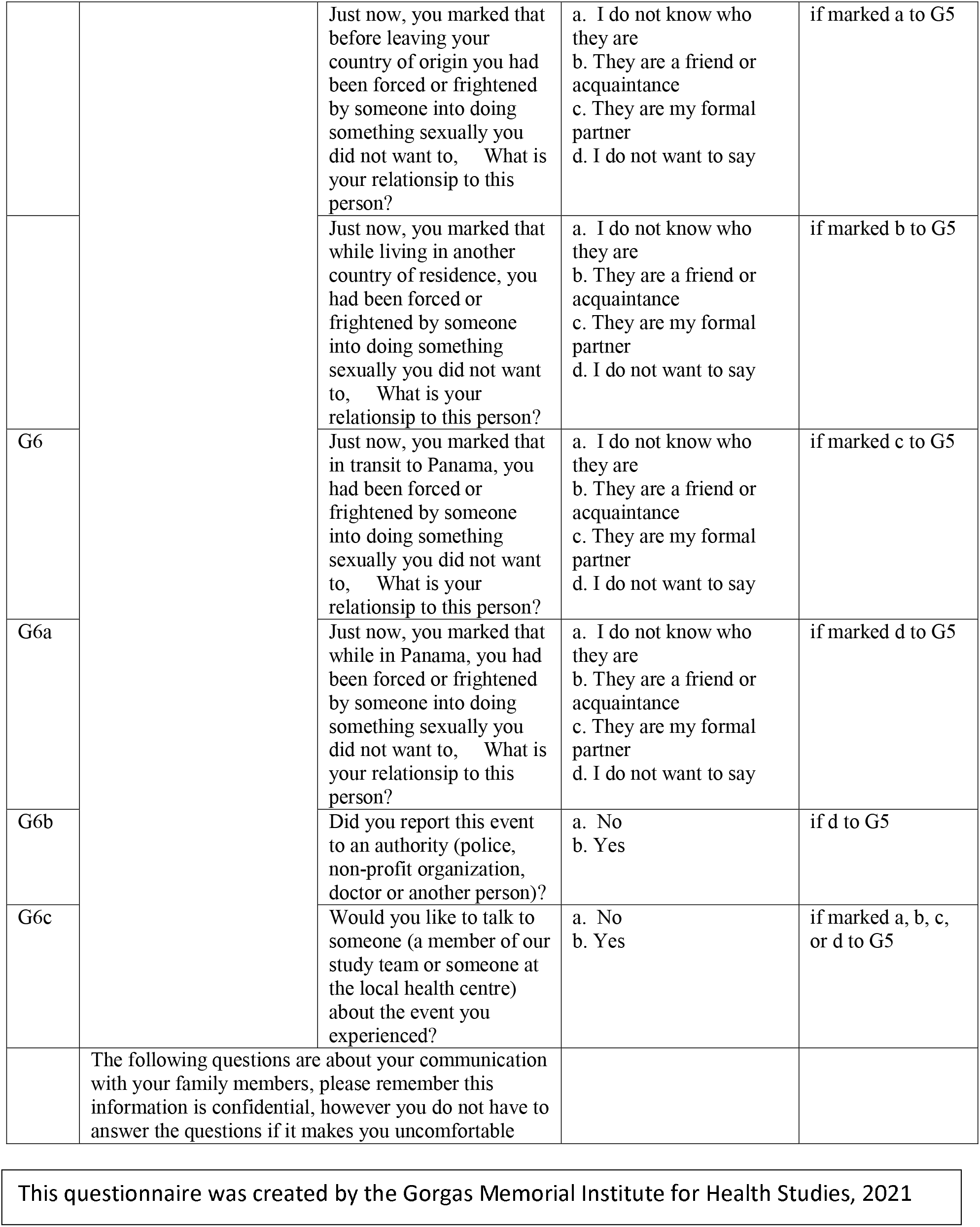
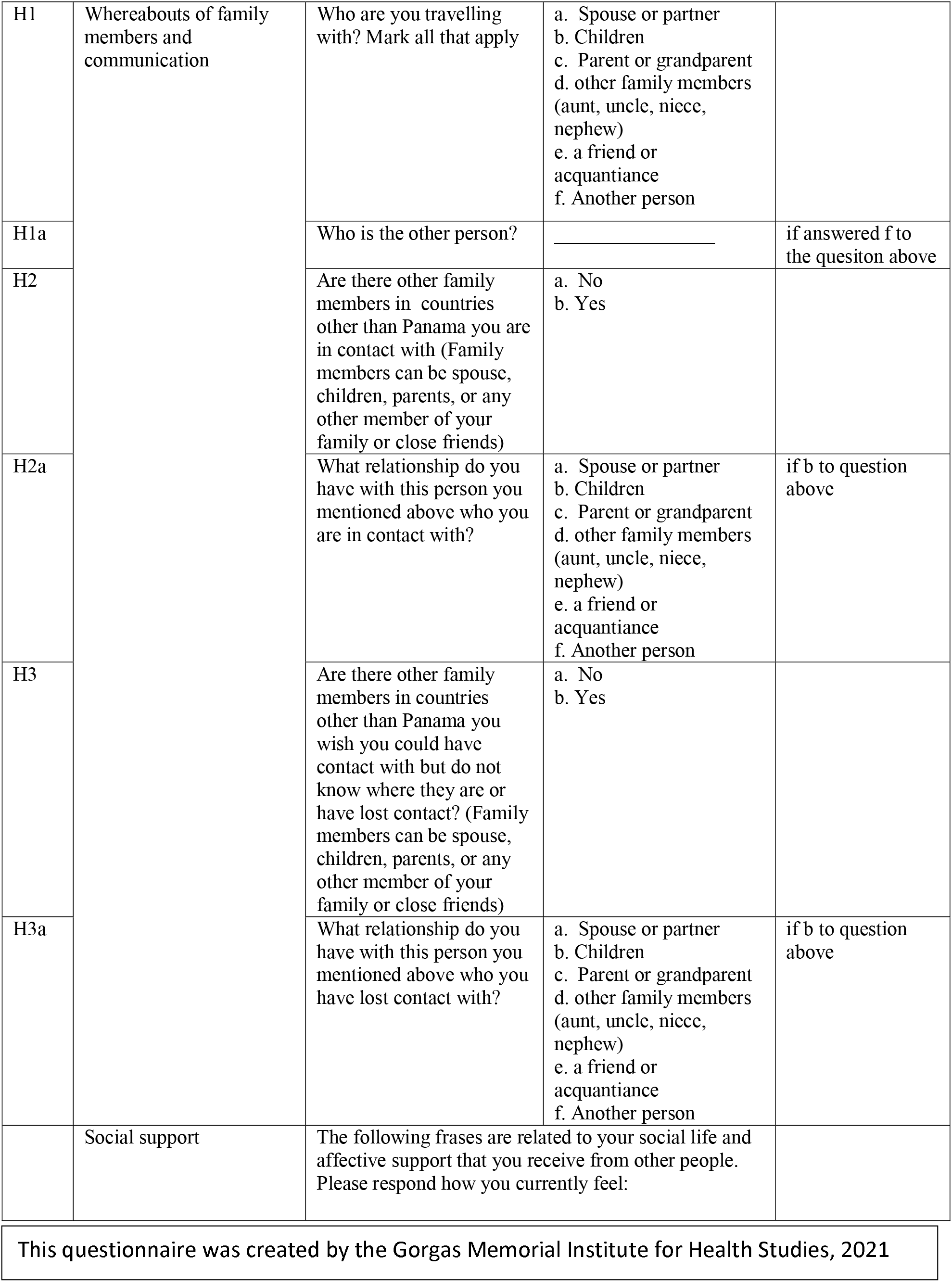
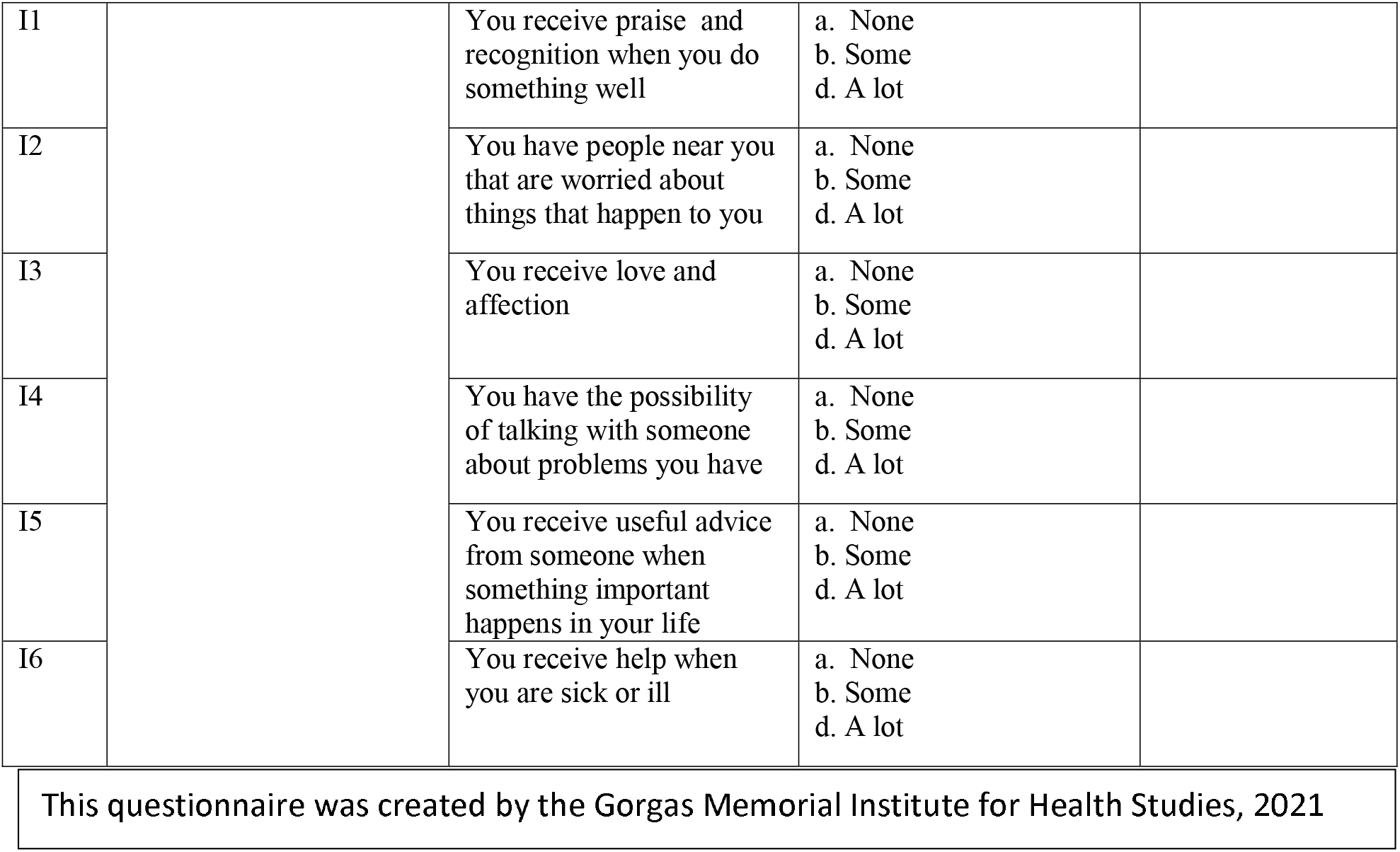

## Supplementary material B: Causes for referral to primary, secondary and tertiary centers.

| **Study component** | **Motive for Referral** |
| --- | --- |
| **Quantitative Questionnaire** |  |
|  | Ongoing distress |
| **Mental health** |  |
|  | Ongoing distress, psychosis and / or risk of self or hetero-aggressive outburst |
| **Sexual and Reproductive Health** |  |
|  | 1. Reported violence: by time since violence occurred and if PEP is possible and support is needed |
|  | 2. HIV positive: refer if had 6 months or more of continuous treatment prior to study and would like to continue treatment; otherwise receive prophylaxis for AIDS-related opportunistic infections. |
|  | 3. Syphilis positive- If congenital syphilis is suspected, if the patient is a kid with <30kg of weight or lactating. |
|  | 4. Pelvic inflammatory disease: Severity symptoms such as fever, nausea, severe abdominal pain; if associated with a pelvic abscess or pregnancy |
| **Pregnancy** | 5. Early pregnancy bleeding (first trimester):<br>5.1 Threatened abortion<br>5.2 Ectopic pregnancy<br>5.3 Gestational trophoblastic disease<br>5.4 Pregnancy loss |
|  | 6. Second and third trimester bleeding:<br>6.1 Placenta previa<br>6.2 Placental abruption<br>6.3 Pregnancy loss |
|  | 7. Complications in the duration of pregnancy<br>7.1 Pre-term labour<br>7.2 Pre-term membrane rupture<br>7.3 Post-term pregnancy |
|  | 8. Hypertensive disorders in pregnancy |
|  | 9. Post-partum hemorrhage |
|  | 10. Infections during pregnancy<br>10.1 Hepatitis B<br>10.2 Toxoplasmosis<br>10.3 Tuberculosis<br>10.4 Genital herpes<br>10.5 HIV<br>10.6 Sepsis |
|  | 11. Mental health disturbances associated to pregnancy:<br>11.1 Postpartum depression<br>11.2 Perinatal dwelling |

| General and Tropical Medicine |  |
| --- | --- |
|  | 7. Neurological symptoms independent of etiology: meningism signs, polyneuropathy, acute flaccid paralysis, convulsions, autonomic symptoms. |
|  | 8. Cardiovascular system: Symptoms associated to myocarditis, endocarditis, pericarditis with clinical suspicion of:<br>8.1 Diphtheria<br>8.2 *Neisseria meningitidis*<br>8.3 *Coxiella burnetii*<br>8.4 *Mycobacterium tuberculosis*<br>8.5 Severe leptospirosis<br>8.6 West Nile river fever<br>8.7 Yellow fever<br>8.8 HIV |
|  | 9. Congestive Heart Failure symptoms that might be due to:<br>9.1 Acute rheumatic fever<br>9.2 Acute Chagas disease<br>9.3 Wet Beriberi<br>9.4 Anemia |
|  | 10. Stroke: by symptoms or clinical suspicion of:<br>10.1 HIV<br>10.2 *Toxoplasma gondii*<br>10.3 Syphilis<br>10.4 Echinococcosis |
|  | 11. Cough by symptoms or clinical suspicion of:<br>11.1 Brucellosis<br>11.2 *Mycobacterium tuberculosis*<br>11.3 *Coxiella burnetii*<br>11.4 *Bordetella pertussis*<br>11.5 Lassa fever<br>11.6 Hantavirus<br>11.7 COVID-19 |
|  | 12. Pneumonia clinically diagnosed or suspicion of the following etiologies:<br>12.1 Aspergillosis<br>12.2 Histoplasmosis<br>12.3 *Burkholderia pseudomallei*<br>12.4 Influenza<br>12.5 *Pneumocystis jirovecii* |
|  | 13. Hemoptysis- Clinical diagnose or suspicion of the following etiologies:<br>13.1 *Yersinia pestis*<br>13.2 *Mycobacterium tuberculosis*<br>13.3 *Mycobacterium abscessus*<br>13.4 *Staphylococcus aureus*<br>13.5 Meliodosis/ *Burkholderia pseudomallei*<br>13.6 *Paragonimus westermanii* |
|  | 14. Pneumonia by vaccine-prevented etiologies |
|  | 15. Hepato-splenomegaly- with high suspicion of the following etiologies:<br>15.1 *Mycobacterium tuberculosis*<br>15.2 Hepatitis virus A,B,C,D,E<br>15.3 Malaria<br>15.4 Visceral Leishmania<br>15.5 Congenital *Trypanosoma cruzi*<br>15.6 *Histoplasma capsulatum* |
|  | 16. Ictericia- with suspicion or diagnosis of the following etiologies:<br>16.1 *Schistosoma japonicum / mansoni*<br>16.2 Malaria<br>16.3 Babesiosis<br>16.4 Hepatitis virus A,B,C,D, E<br>16.5 Dengue virus<br>16.6 Yellow fever<br>16.7 Rift Valley fever<br>16.8 *Coxiella burnetii*<br>16.9 Brucellosis<br>16.10 *Mycobacterium tuberculosis* |
|  | 17. Severe dehydration secondary to diarrhea or dysentery |
|  | 18. Cutaneous affectations secondary to:<br>18.1 *Mycobacterium leprae*<br>18.2 *Mycobacterium tuberculosis*<br>18.3 *Mycobacterium abscessus*<br>18.4 *Mycobacterium ulcerans*<br>18.5 *Corynebacterium diphtheria*<br>18.6 Leishmaniasis<br>18.7 *Neisseria meningitidis*<br>18.8 Ebola/Marburg<br>18.9 Dengue virus<br>18.10 Chikungunya<br>18.11 Rubeola<br>18.12 HIV/Acute Retroviral Symptoms<br>18.13 Measles<br>18.14 West Nile Virus |
| **Nutrition** |  |
|  | Severe acute malnutrition in:<br>-Children from 0 to 6 months<br>-Children from 6 months to 5 years<br>-Children 6-17 years old<br>-18 years or above: Men and women<br>-Pregnant women |

## Supplementary Material C: Distress protocol

The distress protocol will be filled out by the researcher or field assistant involved in the identification of the distress. The following questions will be answered, and if if the response is yes on any, the situation will be reported to the principal investigator.

### Questions to ask of the participant in a state of distress (circle one)

- Did the participant show signs of distress related to a part of the study (partner violence, sexual violence, any other kind of violence or another topic in the study?) **Yes / No**
- Did the participant have to abandon the study due to the distress? Y**es / No**
- Did the participant report a situation of violence (intimate partner or another)? **Yes / No**
- Did or does the participant presenta acute psychosis and or an elevated risk of self o hetero agression? **Yes / No**

**If the participant answered yes to any of these questions, please find the PI or the co-investigator in charge.**

**To be explained to the participant: “we would like to support all participant who present with distress of any kind. Do you have any questions about the study?”**

### Directions after a “yes” was received to the answers above

1. Take the participant to a safe place and ask how they feel
2. Listen to the participant. If they share something, listen with empathy to whatever they want to share.
3. Invite the participant to meet again with you at a specific time, within a couple hours.
4. If the participant comes to the meeting, ask how they are feeling.
5. If the participant is still in a distressed state, or a psychotic break or a situation of increased risk of self or retrogression, they should be taken for an evaluation with the study psychiatrist, who will evaluate if the individual needs further assistance through a referral.

## Supplementary material D: Management of individuals who report sexual violence

If sexual violence has been reported, Ministry of Health authorities will be contacted discreetly; the decisión to contact the authorities will be made by the PI or another co-investigator who is in charge.

The table below indicates where and with what urgency the reference to the Ministry of Health or should be made. These are based on the Ministry of Health (MOH) procedure for sexual violence, rape, incest, and a previously published protocol for children who report violence (*29*).

| **The participant reports** | **urgency of the referral** | **Reported incident** | **Referral to** | **Time to undertake the response and referral** |
| --- | --- | --- | --- | --- |
| Level 1: Forced sexual intercourse, or severe sexual violence in the last 72 hours | Urgent | In the last 72 hours, they have been forced to have sex | Referral and accompaniment to MOH personnel | Within 6 hours of the incident report. |
| Level 2: Severe sexual violence between 3 to 14 days, and minor sexual violence, or if it is more than a week, but the person was in Panamanian territory. | Less urgent, but with notification | Between 3-14 days, the violence occurred. Or, at any time, some less severe act: someone touched their genitals, breasts, or buttocks; forced to touch the genitals, breasts, or buttocks of another person;<br>Other disclosures that do not meet the same criteria for urgency or seriousness as for the urgent need for urgent medical care. | Referral and accompaniment to MOH personnel | As soon as possible (within 24 hours) |
| Level 3: Severe sexual violence in 8 days until the last year, but no incident within Panamanian territory | Without notification, if it occurred outside of Panamanian territory, consult with mental health. | As in level 2, but 8 days until the last year (outside of Panamanian territory). | Go to a mental health consultation, psychology and / or psychiatry staff | When detected |
| No reports of any specific violent act, but the participant indicates that they would like additional support. | Referral to study mental health staff, volunteer |  | Go to a mental health consultation, psychology and / or psychiatry staff | Within 6 hours of having been detected |

## Notes

### Competing Interest Statement

The authors have declared no competing interest.

### Author Declarations

This study has approval from the Comite de Bioetica de la Investigacion del Instituto Conmemorativo Gorgas de Estudios de la Salud (260/CBI/ICGES/21).

### Summary of Updates

This version includes the quantitative questionnaire that will be self-administered.

